# Echinocandin heteroresistance causes prophylaxis failure and facilitates breakthrough *Candida parapsilosis* infection

**DOI:** 10.1101/2022.05.29.22275734

**Authors:** Bing Zhai, Chen Liao, Siddharth Jaggavarapu, Yuanyuan Tang, Thierry Rolling, Yating Ning, Tianshu Sun, Sean A. Bergin, Mergim Gjonbalaj, Edwin Miranda, N. Esther Babady, Oliver Bader, Ying Taur, Geraldine Butler, Li Zhang, Joao B. Xavier, David S. Weiss, Tobias M. Hohl

**Author notes:** Contributed equally (Bing Zhai, Chen Liao, Siddharth Jaggavarapu).

## Abstract

Breakthrough infections of patients on antimicrobial prophylaxis represent a significant and often unexplained cause of morbidity. Here, we reveal that in high-risk patients on micafungin prophylaxis heteroresistance – the presence of a phenotypically unstable, low frequency subpopulation of resistant cells (∼1 in 10,000) – underlies breakthrough bloodstream infections by *Candida parapsilosis* misclassified as susceptible by standard antimicrobial susceptibility testing. By analyzing 219 clinical *C. parapsilosis* isolates from North America, Europe, and Asia, we demonstrate widespread micafungin heteroresistance. To facilitate detection of micafungin heteroresistance, we constructed a predictive machine learning framework that classifies isolates as heteroresistant or susceptible by a maximum of ten genomic features. Our results connect heteroresistance to unexplained prophylaxis failure and demonstrate a proof-of-principle diagnostic approach with the potential to inform clinical decisions.

## Introduction

Antimicrobial resistance leads to over one million annual deaths worldwide and is a severe public health problem ^1^. Heteroresistance is a form of antimicrobial resistance in which a minority subpopulation (sometimes as rare as one in one million cells) of resistant cells co-exists with a majority population of susceptible cells ^2–6^. The minimum inhibitory concentration (MIC) of the resistant cells for a given antimicrobial drug is typically much higher than that of the susceptible cells within the same isolate ^4^. Therefore, treatment with an antimicrobial drug kills susceptible cells but does not kill rare resistant cells, which survive and replicate at concentrations lower than the MIC of the resistant subpopulation. Due to the low frequency of resistant cells, heteroresistance may be undetected by standard antimicrobial susceptibility testing (AST) which typically relies on assays that detect the outgrowth of these cells after 24-48 hours of incubation.

Heteroresistance has been demonstrated to mediate treatment failure in murine infection models ^7–10^, but it has been challenging to determine its role in antimicrobial failure in human infections ^11,12^. Among other confounding variables, many serious infections are treated with multiple antibiotics, which prevents directly linking treatment failure and resistance to a single drug. In contrast, prophylaxis typically relies on administering a single antimicrobial drug, thus providing a simplified scenario to study the impact of drug-resistant phenotypes on antimicrobial failure.

In patients undergoing allogeneic hematopoietic cell transplantation (allo-HCT), randomized prospective clinical trials have established antifungal prophylaxis as the standard of care to prevent fungal infections and to reduce all-cause mortality ^13–16^. Among the three major classes of antifungal drugs, azoles and echinocandins have been approved by the FDA for prophylaxis ^17^. Echinocandin drugs (i.e., micafungin, caspofungin, and anidulafungin) inhibit β-1,3-glucan synthase, which is encoded by the *FKS1* gene and has no mammalian homologs ^18^. Since echinocandins have a favorable toxicity profile, they are commonly used for antifungal prophylaxis in high-risk clinical scenarios.

*Candida parapsilosis* is one of the most common *Candida* species isolated among patients with invasive candidiasis ^19^. Life-threatening *C. parapsilosis* infections caused by fluconazole-resistant isolates in COVID-19 patients have been documented worldwide ^20–23^. *C. parapsilosis* can survive in the gastrointestinal tract as a commensal organism and translocate into the bloodstream during microbial dysbiosis and mucosal injury ^24^. Compared to other *Candida* species, *C. parapsilosis* has higher echinocandin MICs due to a natural P660A polymorphism in the target Fks1 protein ^25^. While classical resistance to echinocandins is rarely observed in this organism ^26,27^, echinocandin prophylaxis or exposure has been linked to an increased frequency of *C. parapsilosis* infections in different medical facilities ^28,29^, with an unclear cause of drug failure.

Here, we describe heteroresistance to micafungin in *Candida parapsilosis* isolates obtained from fecal and blood samples of allo-HCT recipients. Importantly, our results show that patients with intestinal colonization of heteroresistant *C. parapsilosis* isolates have a significantly higher chance of developing breakthrough infections during micafungin prophylaxis, a significant advance in our understanding of the clinical relevance of heteroresistance in prophylaxis failure. Moreover, current antifungal susceptibility tests misclassified heteroresistant *C. parapsilosis* isolates as susceptible, highlighting an urgent need to develop novel diagnostic tools to identify heteroresistance and to inform clinical decision-making. To advance the rapid diagnosis of the heteroresistant phenotype, we employed a large collection of 219 clinical isolates from three continents. We then developed a machine learning framework to predict micafungin heteroresistance from genomic variations. This proof-of-concept approach to identify micafungin heteroresistance in *C. parapsilosis* could help optimize antifungal drug selection, thereby avoiding prophylaxis failure, and improving patient care.

## Results

### *Candida parapsilosis* is a frequent cause of micafungin breakthrough bloodstream infections

To analyze breakthrough fungal bloodstream infections (BSIs) during allo-HCT, we performed a cohort study that included 952 patients who underwent a total of 983 allo-HCTs from January 2016 to December 2020 at Memorial Sloan Kettering Cancer Center (MSKCC) (Extended Data Fig. 1). In the majority of allo-HCTs (n = 966), patients received antifungal prophylaxis with intravenous micafungin (100-150 mg/day) starting seven to ten days prior to allo-HCT for a median of 17 days, prior to switching to triazole-based prophylaxis per institutional practices ^30^. We identified a total of 25 micafungin breakthrough BSIs that occurred within 30 days post transplantation (Extended Data Fig. 1), with a clear predominance of *Candida parapsilosis* as the etiologic agent in these infections (12 out of 25 cases, Fig. 1a).

**Fig. 1.**
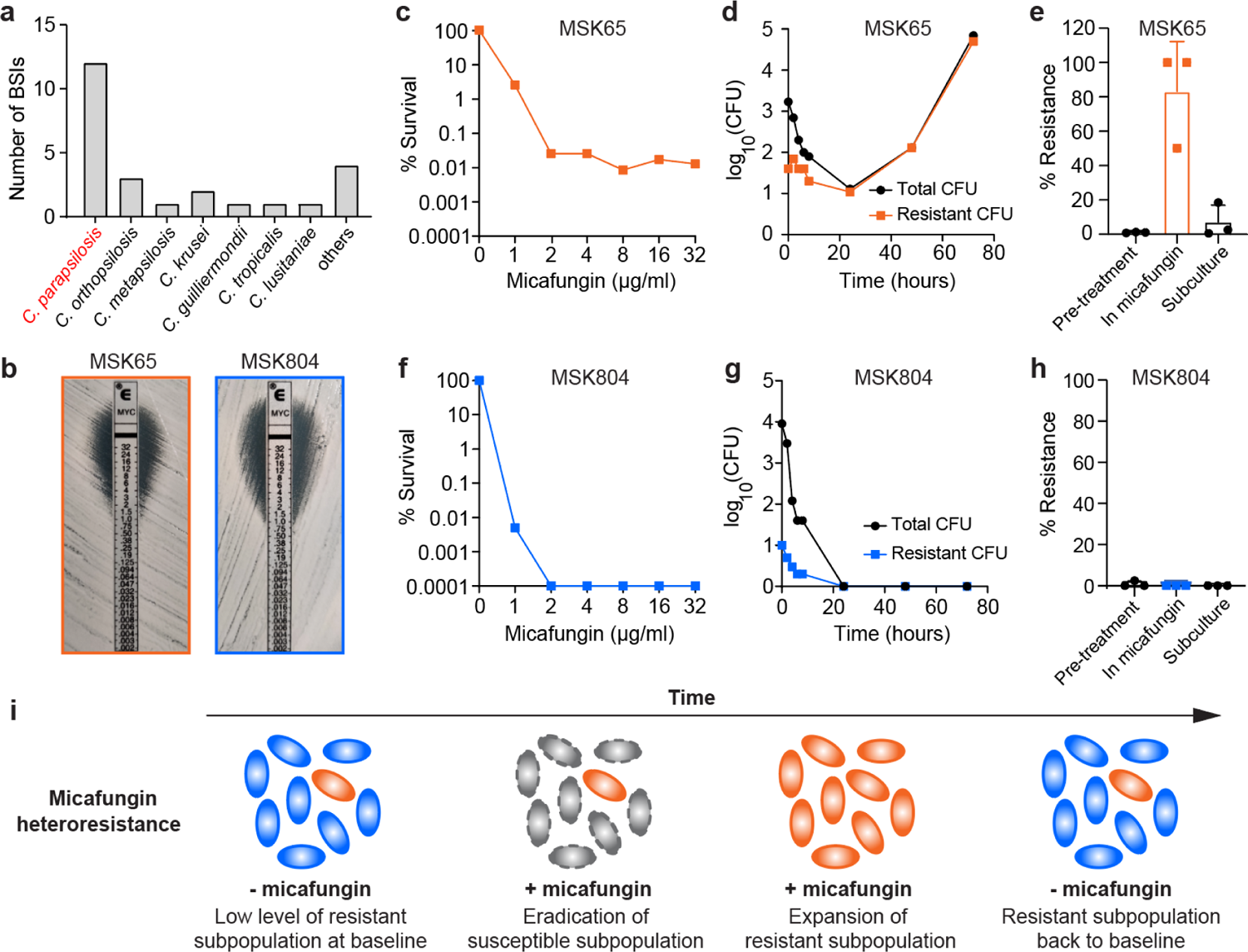
Micafungin heteroresistance in *C. parapsilosis* bloodstream isolates. **(a)** Micafungin breakthrough bloodstream infections (BSIs) in MSKCC allo-HCT recipients between day 0 and 30 post transplantation (between Jan. 2016 and Dec. 2020). The “others” category includes four episodes of breakthrough BSI by *Candida kefyr*, *Trichosporon asahii*, *Capronia munkii*, and *Saccharomyces cerevisiae*. **(b)** Micafungin E-test results of two representative *C. parapsilosis* isolates, MSK65 (heteroresistant) and MSK804 (susceptible). **(c – h)** Population-level responses of MSK65 and MSK804 to micafungin. **(c, f)** Micafungin population analysis profile (PAP) of isolates **(c)** MSK65 and **(f)** MSK804. The % survival was calculated compared to growth on drug-free agar. **(d, g)** CFU dynamics of total and resistant subpopulations of **(d)** MSK65 and **(g)** MSK804 in YPD broth containing 2 µg/mL micafungin. Cultures were plated at indicated time points for enumeration of total (black, circles) and resistant (colored, squares) cells. **(e, h)** Frequency of micafungin-resistant subpopulation of isolates **(e)** MSK65 and **(h)** MSK804 during continuous passage. The frequency was calculated after a 24-hour culture in drug-free YPD broth (Pre-treatment), followed by passaging in YPD broth with 2 µg/mL micafungin (In micafungin) for 48 hours, and a second passage in drug-free YPD broth for 72 hours (Subculture). Bar height: mean; Error bar: standard deviation (n = 3). (i) Graphical illustration of the dynamics of the resistant subpopulation in a micafungin heteroresistant isolate in the presence (+ mica) or absence of micafungin (-mica). Blue cells are susceptible, orange cells are resistant (unstable), and grey cells are those killed by micafungin.

### Micafungin heteroresistance is prevalent in isolates from *C. parapsilosis* breakthrough infections

We were able to retrieve 12 *C. parapsilosis* bloodstream isolates from 10 allo-HCT recipients on micafungin prophylaxis between day 0 and 30 post transplantation (Fig. 1a). In addition, we retrieved three *C. parapsilosis* isolates from three patients who received micafungin prophylaxis after day 30 of transplantation (Extended Data Fig. 1). Notably, all 15 isolates were classified susceptible to micafungin by broth microdilution (Table 1) and E-test (i.e., Epsilometer test; Fig. 1b) assays. However, neither of these conventional ASTs had sufficient sensitivity to determine if there was a low frequency subpopulation of replicating cells resistant to micafungin, i.e., heteroresistance. Thus, we re-analyzed all isolates by the population analysis profile (PAP) assay, the gold standard for detecting heteroresistance ^4^ (see Methods). In PAP assays, 10^6^ fungal cells were plated on solid agar plates that contained micafungin at indicated concentrations. In this study, we defined an isolate as heteroresistant to micafungin when it contained resistant cells with a frequency between 0.0001% (i.e., limit of detection) and 50% across three consecutive serially diluted micafungin concentrations that included 4 µg/ml (2x breakpoint concentration, consistent with the European Committee on Antimicrobial Susceptibility Testing guidelines ^31^, Supplementary Table 1).

As an example, isolate MSK65 was identified as heteroresistant since it harbored a minor subpopulation of micafungin-resistant cells that survived at concentrations up to 32 µg/ml (Fig. 1c). The resistant cells rapidly replicated and became dominant during micafungin exposure (Fig. 1d), indicating that they were not persister cells which cannot replicate exponentially in the presence of an antimicrobial drug ^32^. However, the frequency of resistant cells reverted to the baseline level during subsequent growth in the absence of micafungin, indicating that they were not classical resistant cells (Fig. 1e). In contrast, MSK804 was entirely killed by micafungin exposure without the emergence of a resistant subpopulation (Fig. 1f-h). This isolate was classified as micafungin-susceptible. We confirmed that the heteroresistant phenotypes of blood isolates were consistent across three PAP replicates (Extended Data Fig. 2a and 2b).

Using PAP assays, we found that 11 out of the 15 blood isolates were heteroresistant, with the frequency of resistant subpopulations ranging from 0.008% to 0.8% (Table 1). We further determined the MICs for both the susceptible (MIC-S) and resistant (MIC-R) subpopulations in these 11 heteroresistant isolates (see Methods for definition of MIC-S and MIC-R). Due to the predominance of susceptible cells, the clinical MIC values obtained from broth microdilution aligned well with the respective MIC-S values (Table 1). However, the MIC-R values of these isolates ranged from 32 to 64 µg/ml, indicating an increase of 32- to 64-fold when compared to their corresponding MIC-S values. In addition to micafungin, these isolates were generally, but not uniformly, heteroresistant to two other echinocandin drugs, caspofungin and anidulafungin (Extended Data Fig. 2c and 2d). Together, these findings demonstrate the phenomenon of heteroresistance (HR) to echinocandins in *C. parapsilosis* clinical isolates.

**Table 1.** Micafungin susceptibility profiles of 15 bloodstream *C. parapsilosis* isolates from micafungin breakthrough infections. Clinical minimum inhibitory concentration (MIC) data and susceptibility phenotype were determined by the YeastOne assay. The MIC-S (MIC of the susceptible subpopulation) and MIC-R (MIC of the resistant subpopulation) values were determined by the population analysis profile (PAP) curves (see Methods for details). S, susceptible; HR, heteroresistant; n.d. – not detected; n.a. – not available. The fold difference (Fold diff.) represents the fold change between MIC-S and MIC-R. The resistance frequency (Resistance freq.) indicates the proportion of resistant cells in each isolate that survived at the micafungin breakpoint concentration (2 ug/ml ^31^). Isolates MSK65 and MSK67 were recovered from different blood cultures of patient 1 in Fig. 2c, and isolates MSK485 (HR) and MSK1191 (S) were isolated from different blood cultures of patient 3 in Fig. 2c.

| Isolate | Clinical MIC | Clinical designation | PAP | MIC-S | MIC-R | Fold diff. | Resistance freq. |
| --- | --- | --- | --- | --- | --- | --- | --- |
| MSK65 | 1 | S | HR | 1 | 64 | 64 | 0.033% |
| MSK67 | 1 | S | HR | 1 | 64 | 64 | 0.033% |
| MSK478 | 1 | S | HR | 1 | 64 | 64 | 0.12% |
| MSK485 | 1 | S | HR | 1 | 64 | 64 | 0.075% |
| MSK794 | 2 | S | HR | 1 | 64 | 64 | 0.667% |
| MSK795 | 2 | S | HR | 1 | 64 | 64 | 0.1% |
| MSK802 | 1 | S | HR | 1 | 64 | 64 | 0.8% |
| MSK804 | 1 | S | S | 1 | n.d. | n.a. | n.a. |
| MSK811 | 1 | S | S | 1 | n.d. | n.a. | n.a. |
| MSK812 | 2 | S | HR | 2 | 64 | 32 | 0.375% |
| MSK1191 | 1 | S | S | 1 | n.d. | n.a. | n.a. |
| MSK2384 | 2 | S | HR | 1 | 64 | 64 | 0.014% |
| MSK2386 | 1 | S | S | 1 | n.d. | n.a. | n.a. |
| MSK2387 | 1 | S | HR | 1 | 32 | 32 | 0.04% |
| MSK2390 | 0.5 | S | HR | 1 | 32 | 32 | 0.008% |

**Fig. 2.**
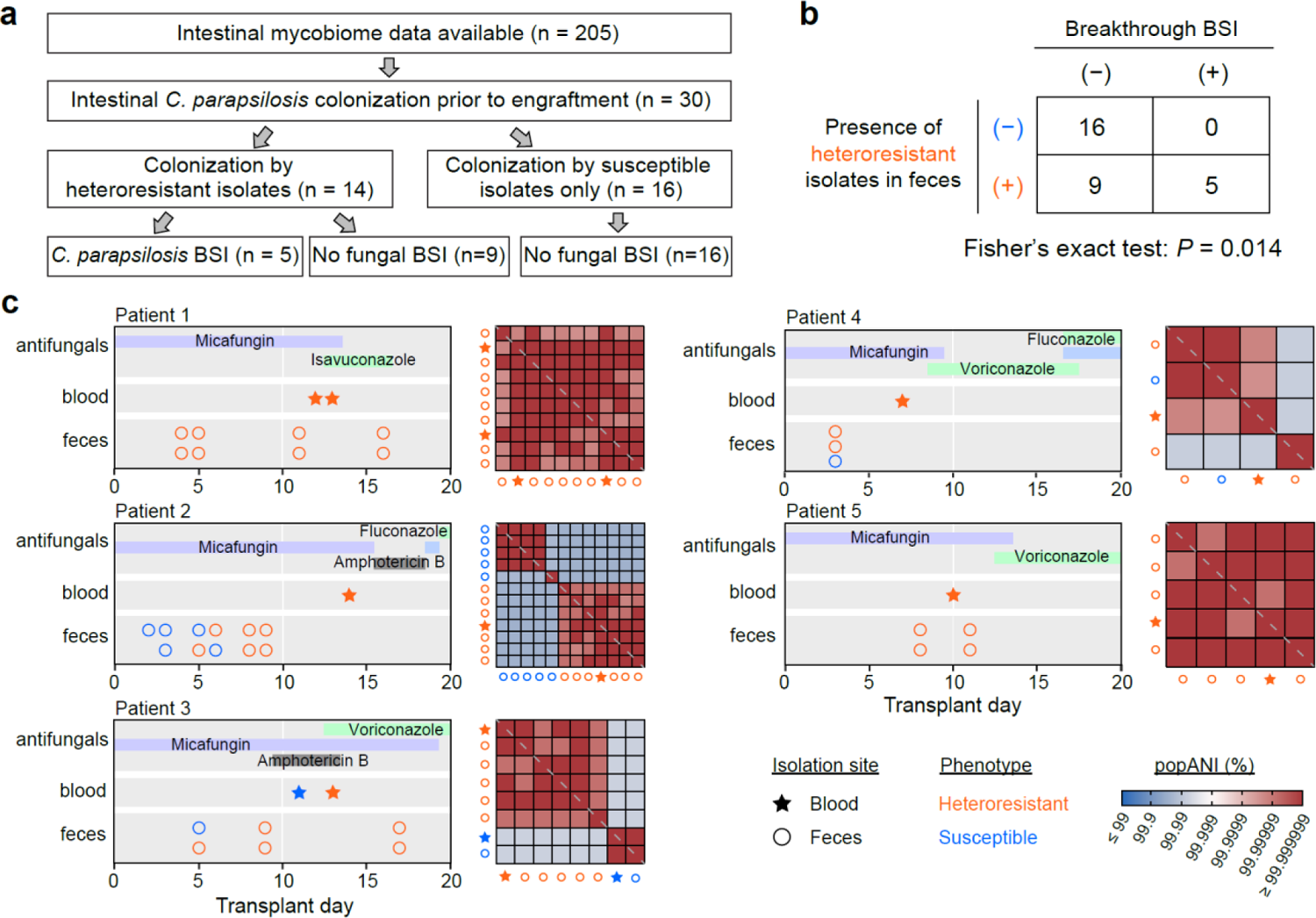
Micafungin heteroresistance and risk of breakthrough *C. parapsilosis* BSIs in allo-HCT recipients. **(a)** A flow chart showing steps involved in determining the number of MSKCC allo-HCT recipients (2016 – 2020) with *C. parapsilosis* intestinal colonization and breakthrough BSIs. **(b)** Fisher’s Exact test (two-sided) on patients with micafungin-heteroresistant *C. parapsilosis* intestinal colonization and breakthrough BSI. **(c)** Comparative genomic analysis between fecal and blood *C. parapsilosis* isolates. Longitudinal medical data are shown to the left for five allo-HCT recipients with *C. parapsilosis* breakthrough BSI, including antifungal drug administration (first row), the time and drug susceptibility of bloodstream isolate(s) (second row), the time of fecal sample collection and the drug susceptibility of corresponding fecal isolate (third row). Pairwise popANI (population average nucleotide identity) between blood (star) and fecal (circle) *C. parapsilosis* isolates were shown in a heatmap to the right of the medical data. At a cutoff of 99.999%, red colors indicate “identical” isolates, and blue colors indicate “distinct” isolates. The grey dashed line on the diagonal indicate self-comparisons between the same isolates (i.e., popANI = 100%). Patient 1 and Patient 2 in this figure were Patient 2 and Patient 3, respectively, in our previous study ^24^.

### Intestinal colonization by a micafungin heteroresistant strain increases the risk of breakthrough bloodstream infection during prophylaxis

A previous study showed that the intestinal tract is a major reservoir for *C. parapsilosis* which can translocate across the intestinal mucosa into the bloodstream and cause BSI in allo-HCT recipients ^24^. To investigate whether micafungin-heteroresistant blood isolates originated from the intestinal tract, we recovered 85 fecal *C. parapsilosis* isolates from 30 patients (5 of whom developed BSI and were investigated in Fig. 1a). Among these isolates, 43 (50.6%) originated from 14 of the patients and were classified as heteroresistant by PAP (Supplementary Table 2). The remaining 16 of the 30 patients were colonized only by susceptible *C. parapsilosis* isolates, and none of these patients developed breakthrough BSI. Among the 14 patients who harbored heteroresistant isolates in their intestinal tract, five patients developed a breakthrough *C. parapsilosis* BSI (Fig. 2a). These data establish an association (P = 0.014; two-sided Fisher’s Exact Test) between intestinal colonization with micafungin-heteroresistant *C. parapsilosis* isolates and ensuing breakthrough BSI (Fig. 2b).

To confirm the gut-to-blood transmission of micafungin-heteroresistant *C. parapsilosis*, we sequenced the genomes of fecal and blood *C. parapsilosis* isolates from the five patients with breakthrough BSIs. In each patient, the bloodstream isolate was identical, defined as >99.999% population average nucleotide identity (popANI) ^33^, to an intestinal isolate collected from the feces prior to BSI, with both isolates having the same heteroresistant or susceptible phenotype (Fig. 2c).

Interestingly, two patients (Patient 2 and 3) had a mixture of heteroresistant and susceptible isolates in their feces before BSI (Fig. 2c). Longitudinal analysis revealed that in Patient 2, the heteroresistant isolates replaced the susceptible isolates, leading to an ensuing BSI by a heteroresistant isolate. In Patient 3, both susceptible and heteroresistant isolates translocated to the bloodstream independently, resulting in two separate episodes of BSI (Fig. 2c). These conclusions remained unchanged using an even more stringent popANI cutoff of 99.9999%.

Taken together, our analysis demonstrates that micafungin-heteroresistant *C. parapsilosis* isolates can colonize the intestine and can subsequently translocate to the bloodstream, thereby presenting a risk factor for the development of breakthrough fungal BSI.

In Patient 2, four of five genomes of susceptible isolates and all genomes of heteroresistant isolates were identical within each category, as defined by the popANI metric. Importantly, the susceptible isolates were distinct from the heteroresistant isolates at the same popANI cutoff (Fig. 2c). Similarly, in Patient 3, two separate clusters of susceptible and heteroresistant isolates were observed. These findings reinforce the notion that the heteroresistance phenotype did not emerge from susceptible isolates during the micafungin prophylaxis period in the patients that we examined. To test this further, we examined 16 patients from whom we isolated at least two identical fecal *C. parapsilosis* isolates at various time points. Micafungin susceptibility phenotypes remained unchanged among these identical isolates in 15 out of 16 patients (Extended Data Fig. 3). In one patient (patient 47), the phenotype of two identical isolates changed from susceptible to heteroresistance, due to a two-fold change in the MIC of the resistant subpopulation in the later isolate.

### Association between micafungin heteroresistance phenotype and genomic variations in clinical isolates from three continents

To expand the scope of the study, we included 118 *C. parapsilosis* clinical isolates from MSKCC (see Methods) and supplemented the collection with an additional 18 isolates from other hospitals in the United States, 73 isolates from 32 different hospitals from 20 provinces in China, 9 isolates from Germany (Göttingen), and one isolate from France (Nantes), yielding a total of 219 isolates (Supplementary Table 2). Using the PAP method, we found the presence of micafungin-heteroresistant *C. parapsilosis* isolates in hospitals from all three continents (68/136, 11/73, 7/10 micafungin-heteroresistant isolates from the North America, Asia, and Europe, respectively). This finding highlights the global prevalence of this emerging and clinically significant phenotype.

To understand the genomic underpinnings of micafungin heteroresistance, we sequenced the genomes of these 219 isolates and profiled the genomic variations (GATK, see Methods).

Chromosome-level variations such as aneuploidy have been reported in antifungal heteroresistance in *C. parapsilosis* and other human fungal pathogens^34–36^. We identified 12 out of the 219 isolates (5.5%, eight heteroresistant and four susceptible isolates) with different patterns of trisomy (seven, four, and one isolates have an extra copy of chromosome VI, III, and V, respectively, see Extended Data Fig. 4). Although 8 out of 12 isolates with aneuploidy were micafungin-heteroresistant, the study may lack power to confirm or reject a correlation (two-sided Fisher’s exact test, P = 0.066). An even larger collection of *C. parapsilosis* isolates would help determine further a relationship between aneuploidy and micafungin heteroresistance. Notably, most heteroresistant isolates (78 of 86) did not exhibit aneuploidy.

We next characterized copy number variations (CNVs), which have been linked with azole tolerance ^37^, across the 219 *C. parapsilosis* genomes. We refined a previous algorithm on CNV characterization ^38^ and defined the copy number of each ORF by two features (see Methods and Extended Data Fig. 5 for illustrative examples): the first feature describes whether an ORF presents a full or a partial amplification or deletion (CNV_categorical or CNV_cat), and the second feature quantifies the copy number of amplified or deleted regions (CNV_quantitative or CNV_quant). With this approach, we identified 3,317 ORFs that contain amplified or deleted regions in at least one isolate in this dataset (Supplementary Table 3). Among the 2,774 ORFs with amplifications, only 53 show overlaps with inverted repeats of at least 100 bp, suggesting that these CNVs were unlikely to be technical artifacts (Supplementary Table 4).

We identified 43,531 single nucleotide variants (SNVs) and 3,722 small insertion/deletions (INDELs) among the 219 isolates (Supplementary Table 5). These variations served as the basis for constructing a phylogenetic tree (see Methods; Fig. 3). Notably, the majority of heteroresistant isolates cluster within one clade, establishing a robust correlation between the heteroresistance phenotype and phylogeny (Moran’s I = 0.30, P < 0.0001).

**Fig. 3.**
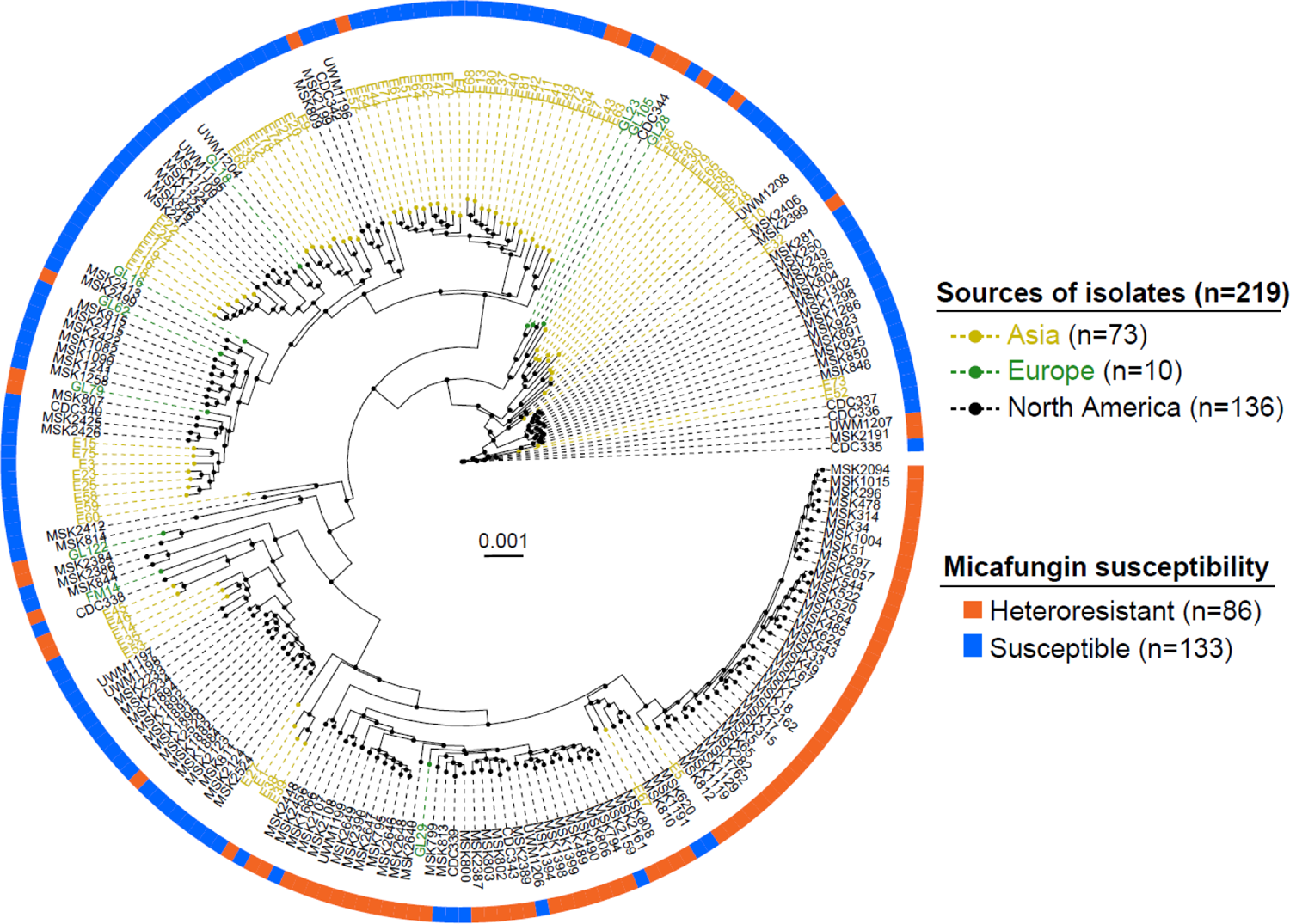
Phylogenetic tree of 219 *C. parapsilosis* isolates collected from three continents. The outer circle indicates the micafungin susceptibility phenotype of each isolate.

We next sought to identify variations that are associated with micafungin heteroresistance among the profiled SNVs, INDELs, and CNVs. We first encoded these variations as a mixture of continuous (CNV_quant) and categorical (SNVs, INDELs, CNV_cat) features and then merged redundant features if they correlated perfectly with each other across all 219 genomes (Methods). This resulted in a non-redundant set of 6,806 features (Supplementary Table 6). Using logistic regression (see Methods), we identified 453 features that were significantly associated with the heteroresistance phenotype after multi-test correction (Fig. 4a, Supplementary Table 7). We confirmed that the top five features with both positive and negative associations did not overlap with the inverted repeats identified above (Supplementary Table 4). The most significant feature (Feature 4376) was a homozygous alternative allele at the 5’UTR of CPAR2_206130 (P_adj._ = 2.4e-15). The *Saccharomyces cerevisiae* ortholog of this gene encodes a microtubule-associated protein that plays a role in fungal cell wall organization ^39^ (Fig. 4b). The second most significant feature (Feature 2154) was the full deletion of CPAR2_600440 (P = 2.9e-15), which encodes a putative glycosylphosphatidylinositol (GPI)-anchored protein involved in cell wall integrity and cell-cell interactions ^40^ (Fig. 4b). Both features remained the top-ranking ones when we examined the associations by LASSO (Least Absolute Shrinkage and Selection Operator; see Methods and Supplementary Table 7).

**Fig. 4.**
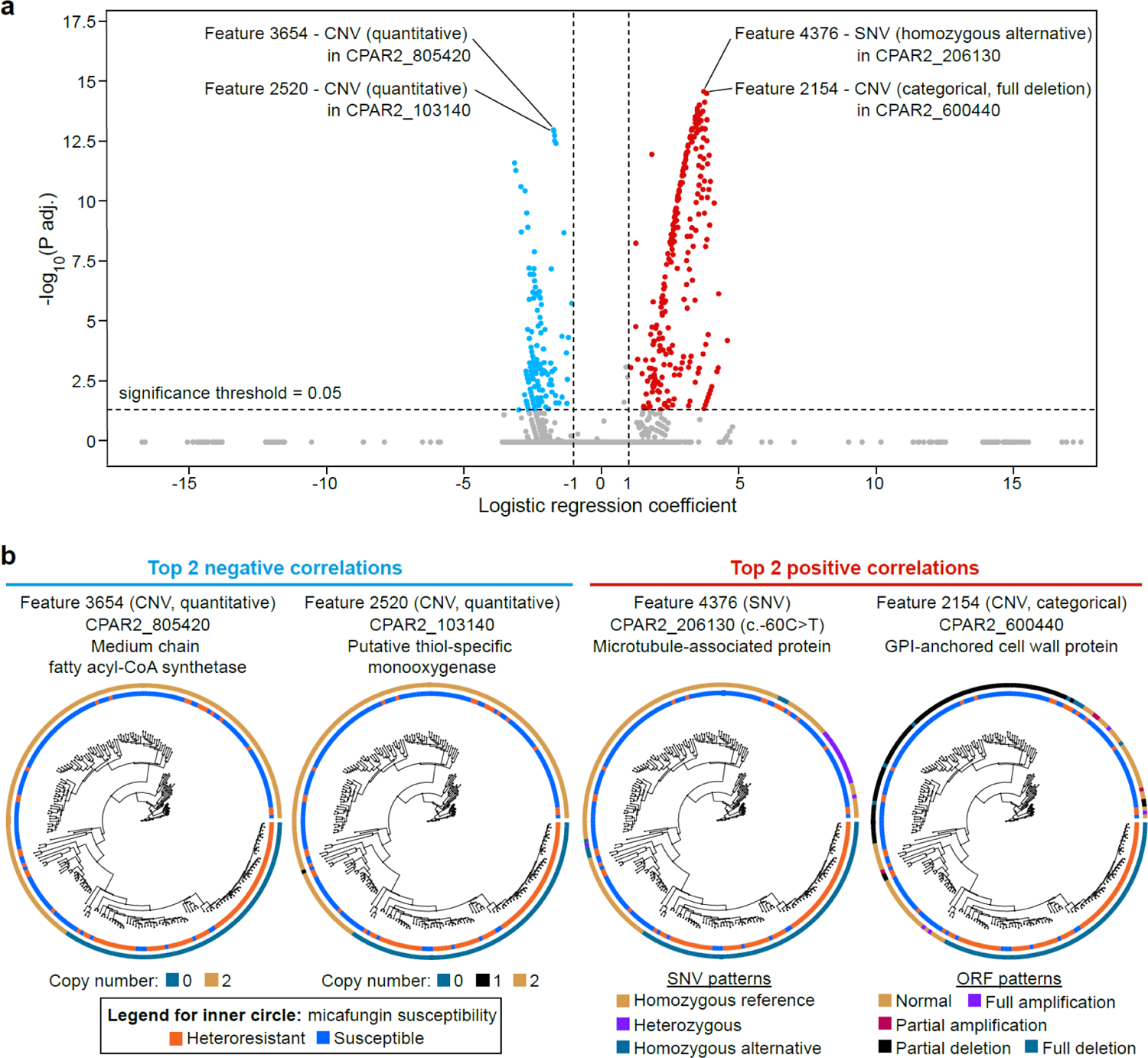
Top *C. parapsilosis* genomic features associated with micafungin heteroresistance. **(a)** Volcano plot of 6,806 *C. parapsilosis* genomic features. **(b)** The top two features positively and negatively associated with micafungin heteroresistance and the annotations of the open reading frames that contain the four features. Distribution of the features were plotted in alignment with the phylogenetic tree. Chromosome locations: Feature 4376: Chr 2; Feature 2154: Chr 6; Feature 3654: Chr 8; Feature 2520: Chr 1. SNV, single nucleotide variation; CNV, copy number variation.

All 453 features with a significant correlation with heteroresistance also correlated with phylogeny (Moran’s I test, P_adj._ < 0.05). Among these features, Feature 100 (a homozygous alternative allele within CPAR2_109680, P_adj._ = 5.7e-10) was the only one found in an orthologue of the gene encoding the primary drug target. However, this correlation could be due to phylogeny (Extended Data Fig. 6a). We applied a phylogenetic LASSO model to re-evaluate the associations of genomic features with heteroresistance while controlling for their phylogenetic dependences (see Methods). This approach yielded 40 features with non-zero coefficients, where a homozygous alternative variant (Feature 4470) in CPAR2_206800 had the largest positive coefficient (Supplementary Table 8). Although this feature did not completely overlap with the heteroresistant isolates outside the major cluster, it successfully identified the heteroresistance phenotype of three isolates within a phylogenetic region that is dominated by susceptible isolates (Extended Data Fig. 6b). Taken together, the phenotype of micafungin heteroresistance in *C. parapsilosis* is associated with genomic variations and these associations are strongly, but not exclusively, coupled with phylogeny.

### Machine-learning-based diagnosis of micafungin heteroresistance

Given the clinical significance of micafungin heteroresistance, a rapid diagnosis with high accuracy is critical for the rapid administration of effective antifungal drugs. However, current laboratory approaches (i.e., PAP assay) require incubation times up to several days with high material and labor demands. To facilitate the diagnostic process, we developed a machine learning framework (Extended Data Fig. 7) that predicts the micafungin heteroresistance phenotype from the profiled genomic features found in the 219 clinical isolates. Given that multiple isolates from a single patient may share identical genomes (i.e., popANI ≥ 99.999%), separating these isolates into the training and test sets could lead to data leakage and compromise the model evaluation. To address this issue, we removed the duplicates of identical isolates collected from each patient (see Methods, Extended Data Fig. 8), resulting in a total of 160 non-redundant isolates for further analysis. The machine learning framework includes feature standardization, feature selection, hyperparameter optimization, and classification.

In the machine learning framework, we assessed three feature selection methods (logistic regression, LASSO, a neural-network-based approach ENNS ^41^) along with three tree-based classifiers: Decision Tree (DT), Random Forest (RF), and Extreme Gradient Boosting (XGBoost). Our approach limited each feature selection method to choose a maximum of ten features to mitigate the risk of overfitting and, more importantly, to reduce the time and effort required to measure the selected features in the laboratory (see Discussion). All model hyperparameters, including the number of features selected, were optimized through a grid search in a 5-fold cross-validation setting. The 160 non-redundant isolates (Supplementary Table 9) were divided into a training set (80%) and a test set (20%), the process of which was repeated 50 times. On average, the combination of logistic regression and XGBoost consistently outperformed the other eight combinations in F1 score (Fig. 5a) and other evaluation metrics (precision, recall, and accuracy, Extended Data Fig. 9a). This combination achieved a median precision of 0.78, a recall of 0.70, an F1 score of 0.74, and an accuracy of 0.84. This strategy can be further simplified by developing SNV-only models, which have equivalent performance to the models developed with SNVs, INDELs and CNVs (Fig. 5a). To confirm that the optimal model captures more than spurious correlations, we randomly permuted the heteroresistance labels to simulate the shuffled data and retrained the model. As expected, the resulting median F1 score never exceeded 0.25 across three repetitions (Fig. 5b). This striking difference strongly indicated that our model effectively captured meaningful genomic features of micafungin heteroresistance.

**Fig. 5:**
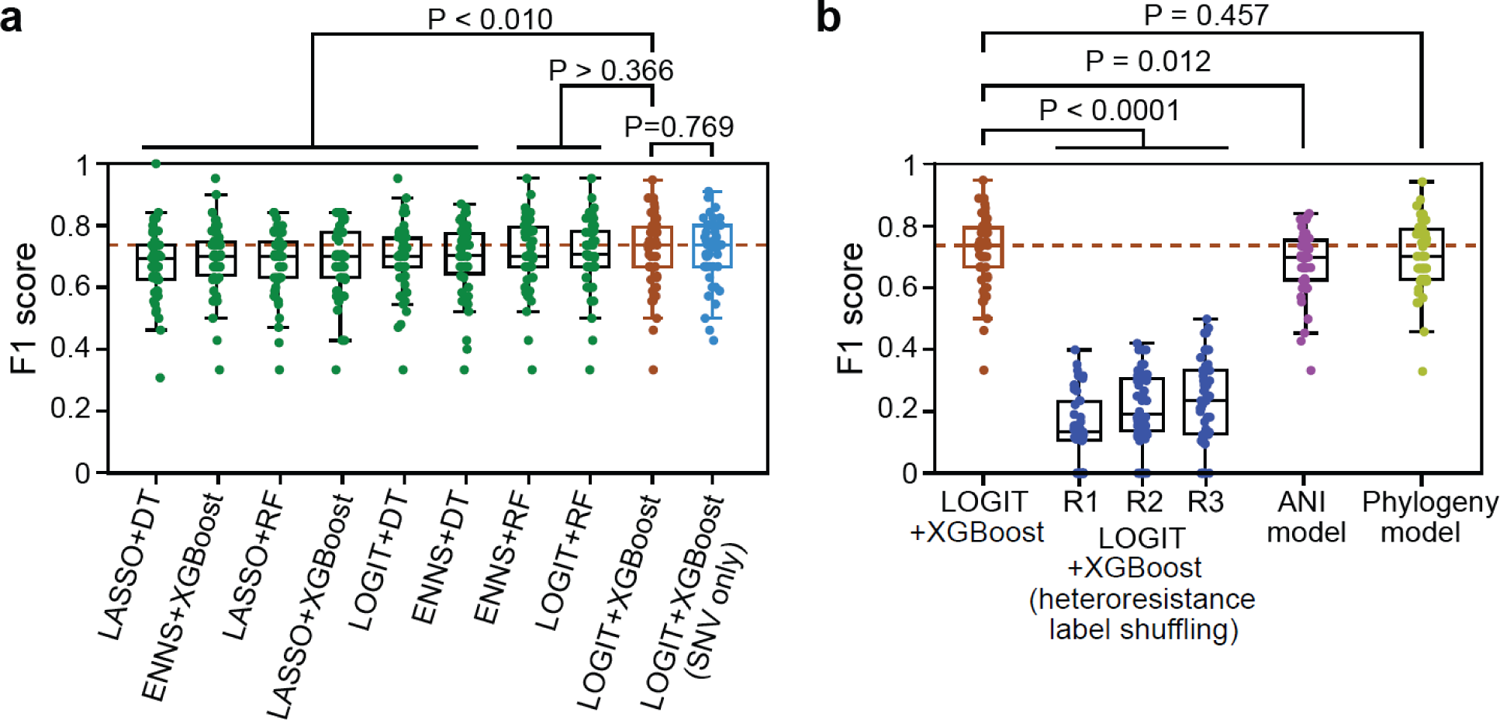
Evaluation of machine learning model performance in predicting the micafungin heteroresistance phenotype. F1 score is an evaluation metric that combines precision and recall. Unless specified otherwise, all models selected features from a combined pool of SNV (single nucleotide variant), INDEL (insertion/deletion), and CNV (copy number variation) features. In all boxplots, each dot represents a single train-test split, which was randomly repeated 50 times. **(a)** Machine learning models constructed by choosing among three feature selection algorithms (LOGIT, Logistic regression; LASSO, Least Absolute Shrinkage and Selection Operator; ENNS, Ensemble Neural Network Selection) and three classifiers (DT, Decision Tree; RF, Random Forest; XGBoost, Extreme Gradient Boosting). **(b)** Comparison of the best-performing combination (LOGIT+XGBoost) with alterative models. R1, R2, and R3 represent three repeats of retrained LOGIT+XGBoost models using randomly shuffled heteroresistance data. The ANI (average nucleotide identity)-based model quantified genomic similarity between isolates by popANI and assigned a test isolate the same heteroresistance label of its genomically closest isolate in the training set. Phylogeny-based model placed test isolates into a reconstructed phylogeny of training isolates and then performed hidden-state prediction. Two-sided paired-sample *t*-test: exact P values are provided.

Given the strong association between heteroresistance and phylogeny (Fig. 3), we developed an alternative phylogeny-based predictive approach (see Methods). This approach placed genome sequences of the test isolates into a reference phylogenetic tree created by the training isolates and conducted hidden-state prediction to infer their micafungin heteroresistance labels. Interestingly, the best machine learning model achieved a slightly higher median F1 score (0.74) than the phylogeny-based approach (0.71, Fig. 5b). Although the difference in performance did not attain statistical significance (paired *t*-test, P = 0.457), both methods substantially outperformed a popANI-based method (Fig. 5b), which simply assigned a test isolate the same heteroresistance label as its most similar (i.e., highest popANI) isolate in the training set. These findings indicate that the features chosen by the machine learning model exploit the phylogenetic signal of heteroresistance and expand on it to achieve predictive power. The most commonly misclassified isolates, including false positive and false negative predictions (Extended Data Fig. 9b), showed micafungin susceptibility phenotypes inconsistent with their phylogenetic neighbors. Collectively, our proof-of-concept machine learning framework demonstrated its effectiveness in predicting the micafungin heteroresistant phenotype based on a small set of genomic features. This innovative approach could facilitate the identification of antimicrobial heteroresistance in clinical settings.

## Discussion

This study shows that micafungin heteroresistance promotes breakthrough *C. parapsilosis* infections in allo-HCT patients on micafungin prophylaxis. While both heteroresistant and susceptible *C. parapsilosis* isolates could colonize the intestinal tract, micafungin heteroresistant isolates were enriched in patients who developed breakthrough BSI. Factors such as host immunity or gut microbiota composition may also contribute to fungal translocation from the gut to the bloodstream. Nonetheless, our results show intestinal colonization by a micafungin heteroresistant strain increases the risk of developing breakthrough BSI. Importantly, we did not observe any instances of classical, stable resistance to micafungin in this study. These findings highlight heteroresistance, rather than classical resistance, as a major cause of breakthrough *C. parapsilosis* infections during echinocandin prophylaxis.

Identifying heteroresistance is challenging in the clinic because the resistant subpopulation reverts to a very low frequency after removing the drug (i.e., the initial blood culture process and the ensuing subculture of fungal cells for drug susceptibility testing). Standard clinical ASTs fail to detect micafungin heteroresistance. PAP assay – the gold standard for heteroresistance in research laboratories – is laborious and can take days to weeks to identify slow-growing resistant cells. Isolate phylogeny, which we found to be a robust predictor of micafungin heteroresistance in *C. parapsilosis*, requires whole genome sequencing which is also time-consuming and computationally involved. In contrast, the machine learning models introduced in this study include at most ten genomic features, which can be rapidly measured by regular Sanger sequencing for SNVs and INDELs, quantitative PCR for CNVs. This approach can bypass the time needed to culture fungal isolates during AST testing and is more precise than phylogeny-based prediction (Fig. 5). Due to the increasing prevalence of antimicrobial resistance, this proof-of-concept approach may advance current diagnostics and support rapid, personalized clinical decisions in therapy and infection control. A future research direction is to harness this strategy to detect intestinal colonization by heteroresistant *C. parapsilosis* using metagenomic data.

Predicting micafungin heteroresistant phenotype can be improved in future studies by using a larger, more diversified collection of *C. parapsilosis* isolates combined with their micafungin susceptibility phenotypes. Classical resistance or tolerance to echinocandin drugs has been attributed to chromosomal mutations in the *FKS* or *MSH2* genes in *Candida* species ^25,42^.

Here, with the exception of Feature 100 (SNP in CPAR2_109680, Extended Data Fig. 6a), we did not identify variations in *FKS* or *MSH2* genes associated with micafungin heteroresistance. In fact, insights from azole tolerance studies have indicated that, unlike conventional resistance which often arises from mutations of the direct drug target, tolerance or heteroresistance may hinge on changes in stress response pathways ^2^. Elucidation of the biological functions of these genes and the implications of the identified variations require further investigation, and could be guided by future models trained on larger datasets. Future epidemiologic studies should also clarify the potential spread of heteroresistant strains within hospital or healthcare settings.

Finally, we examined the presence of echinocandin heteroresistance in another medically relevant fungal pathogen, *Candida auris*. *C. auris* are highly drug-resistant pathogenic fungi that have rapidly spread across the world since their identification in 2009. More than 99% of the *C. auris* isolates in the United States are resistant to fluconazole, 50-60% are resistant to amphotericin B, and less than 10% are resistant to echinocandins ^43–45^. Thus, echinocandin drugs are the first-line empiric therapy for *C. auris* infections, as recommended by the CDC ^46^. We identified micafungin heteroresistance among *C. auris* isolates from the CDC/FDA AR Bank using PAP analysis (Extended Data Fig. 10). Interestingly, recent reports describe echinocandin treatment failures caused by susceptible *C. auris* as classified by standard AST, along with emergence of resistance while the patients were being treated with echinocandins ^43,44^. It is worthwhile to determine if some of these isolates are heteroresistant to echinocandins, which may link heteroresistance to these reported treatment failures. Such potential treatment failures, together with the prophylaxis failures that we observed in allo-HCT recipients, would highlight further the urgent need for rapid detection of echinocandin heteroresistance in clinical settings.

In summary, our study has uncovered the existence of heteroresistance to echinocandin drugs in *Candida* species and has linked this phenotype to prophylaxis failures in allo-HCT recipients. The proof-of-concept machine learning framework formulated here lays the foundation for future development of rapid and precise diagnostic methods for heteroresistance.

## Methods

### Inclusion criteria for allo-HCT recipients and *Candida parapsilosis* isolates at MSKCC

Patients that underwent allo-HCT at Memorial Sloan Kettering Cancer Center (MSKCC) between Jan. 1^st^, 2016 to Dec. 31^st^, 2020 were examined for micafungin breakthrough fungal bloodstream infections. In this study, we defined micafungin breakthrough bloodstream fungal infections as a positive fungal blood culture if a patient received a minimum of one day of micafungin prophylaxis prior to the blood culture positivity, or if micafungin prophylaxis was discontinued within three days before the blood culture positivity. 15 isolates were retrieved from documented breakthrough BSI in these patients: 12 isolates from 10 patients who developed BSI between transplant day 0 to 30 and three isolates from three patients who developed BSI after day 30. The medical record of non-allo-HCT patients with *Candida parapsilosis* bloodstream infections from the same study period (2016 to 2020) were also reviewed (Supplementary Table 2). A total of 14 bloodstream *C. parapsilosis* isolates from these non-allo-HCT patients who developed *C. parapsilosis* BSI based on the criterion of having a positive blood culture were obtained from clinical micro lab at MSKCC.

Fecal biospecimens from allo-HCT recipients were drawn from a prospective fecal collection biobank, which has been described previously ^47^ (MSKCC IRB protocol 16-834). Fecal samples from a subset of 205 patients (collected from −10 to 30 day of transplantation) had been cultured for viable fungal isolates as previously described ^24^. Among them, a total of 30 patients were confirmed with intestinal colonization by *C. parapsilosis* from fecal culture. For the five patients with intestinal colonization of *C. parapsilosis* before BSI, at least one isolate from each fecal sample was randomly picked for further analyses. For the remaining 25 patients with intestinal colonization but no fungemia, one to two *C. parapsilosis* isolates from each *C. parapsilosis* culture positive fecal samples were randomly picked. Detailed fecal isolate – patient information data are listed in Supplementary Table 2. Fecal biospecimens from eight healthy donors ^48^ were also examined for the *C. parapsilosis* culture positivity. Two fecal samples were identified as positive and two *C. parapsilosis* isolates from each sample were randomly picked. Taken together, there were a total of 89 fecal *C. parapsilosis* isolates from MSKCC included in this study. Detailed information of patient demographics, antimicrobial exposure, bloodstream infections, and fecal isolates are available in Supplementary Table 2.

### *C. parapsilosis* isolates from non-MSKCC hospitals

Among the 18 *C. parapsilosis* isolates from United States other than MSKCC, nine isolates were acquired from David Andes’s lab at University of Wisconsin-Madison. The other nine isolates were acquired from the CDC/FDA AR Bank. 73 Asian isolates were acquired from China Hospital Invasive Fungal Surveillance Net (CHIF-NET). These isolates were collected from 32 hospitals from 20 different provinces in China. Ten European isolates include one from Nantes, France and nine from Göttingen, Germany. The detailed information of the isolates are available in Supplementary Table 2.

### Classification of micafungin heteroresistance

In this study, an isolate was classified as heteroresistant if the frequency of resistant cells is at least 0.0001%, but remains lower than 50%, across all three consecutive serial-diluted concentrations which include 4 µg/ml (2x breakpoint concentration, EUCAST ^31^). The possible set of three consecutive concentrations could be 1, 2, and 4 µg/ml, or 2, 4, and 8 µg/ml, or 4, 8, and 16 µg/ml. Additionally, we determined the MIC-S (MIC of the susceptible subpopulation) and MIC-R (MIC of the resistant subpopulation) from PAP curves. MIC-S was defined as the minimum concentration at which survival subpopulation drops below 50%. On the other hand, MIC-R was defined as the minimum concentration at which the survival subpopulation falls below the detection limit (0.0001%) at this concentration and all other concentrations beyond it.

### Population analysis profile assay (PAP)

YPD agar (Sigma) plates were made with each antifungal drug at the indicated concentrations. The stocks of micafungin (Biovision), caspofungin (Biovision) and anidulafungin (Biovision) were made by dissolving each drug in molecular biology grade water (Corning). Overnight cultures of all the isolates were grown in 2 mL of YPD broth (RPI) in 8 mL round-bottom tubes (Corning) at 35°C with 200 rpm shaking. Ten-fold serial dilutions of the overnight cultures were made in PBS in 96-well plates (Corning). 5 µL of each dilution, from undiluted to 10^-6^, were spotted on the agar plates. Plates were incubated for 48 to 72 hours at 35°C before CFUs were enumerated. To calculate the percentage survival, CFUs from the plates containing antifungals were divided by those from the plates without antifungals.

### Time-kill assays

2 mL of YPD broth supplemented with 2 µg/mL micafungin for *C. parapsilosis* or 0.5 µg/mL micafungin for *C. auris*, were inoculated with 10^6^ CFU from an overnight culture. These YPD cultures with micafungin were incubated at 35°C with shaking at 200 rpm for up to 72 hours. At each indicated time points, 50 µL aliquots were acquired from the cultures and CFUs were counted by plating serial dilutions on plates with and without micafungin at the same concentrations as the respective broth cultures.

### Antifungal resistance stability assay

For antifungal treatments, *C. parapsilosis* cultures were sub-cultured (1:100) from a drug-free overnight culture into YPD with 2 µg/mL micafungin and grown at 35°C with 200 rpm shaking for 48 hours. Micafungin-free subcultures were performed by diluting the micafungin culture 1:1,000 into fresh YPD media and the cultures were grown for 72 hours. Cultures were diluted in PBS and plated on YPD-agar plates with and without micafungin to determine the proportion of resistant *C. parapsilosis* cells.

### E-tests

Micafungin E-test strips were obtained from Biomerieux (Marcy-l’Étoile, France). The E-tests were performed according to the manufacturer’s instructions. Briefly, 5 colonies from an overnight YPD agar plate were dissolved into sterile PBS. The OD was adjusted to 0.5 McFarland standard. The solution was applied to RPMI 1640 plates with MOPS and 2% Glucose (Remel) with a cotton swab. The E-test strip was laid on the plate and the images were captured after 48-hour incubation at 35°C.

### Average nucleotide identity (ANI) and identity between isolates

ANI is a quantitative metric for comparing the genomic similarity between two microbial genomes. We calculated the pairwise population ANI (popANI) values for all 219 *C. parapsilosis* isolates by the inStrain package ^33^ (version 1.6.3). A recommended threshold of 99.999% was used to determine whether two isolates are identical or distinct. However, pairwise comparisons may pose challenges in identifying identical isolates when more than two isolates are compared. For example, the identity between isolate A and C cannot be determined when isolate A and B are identical (popANI ≥ 99.999%) and isolate B and C are identical (popANI ≥ 99.999%), but isolate A and C are not identical (popANI < 99.999%).

To address this issue, we developed a computational algorithm to extract the largest clusters in which the popANI value between any two isolates is at least 99.999% and no additional isolate has popANI ≥ 99.999% with all isolates in the cluster. Within each such cluster, all isolates are identical to each other. To identify these clusters, we first conducted hierarchical clustering of the binarized popANI matrix at the 99.999% threshold. Using the “fcluster” function in the Python SciPy package (version 1.10.1), we systematically reduced the maximum allowed inter-cluster distance until the presence of a cluster where all pairwise popANI values among its isolates are at least 99.999%. If multiple such clusters exist, we selected the largest cluster and assigned it a cluster ID. Subsequently, isolates in this cluster were removed from the popANI matrix, and hierarchical clustering was repeated on the remaining isolates. We iterated these steps until no clusters containing more than one isolate could be identified. The remaining isolates were each treated an autonomous cluster. By dividing all isolates into clusters, we define that any two isolates are identical if they belong to the same cluster; otherwise, they are distinct.

### Genomic variation calling, filtering, and annotation

Reads were mapped against *C. parapsilosis* CDC317 reference genome ^49^ using BWA-mem (version 0.7.17-r1188) with parameters -M -R ‘RG’, where ‘RG’ represents read group information generated for each isolate. Duplicate reads were marked using the GATK (version 4.1.9.0) MarkDuplicates tool as per GATK best practices. SNVs and INDELs were identified using the HaplotypeCaller tool from GATK. The resulted GVCF files were combined using the CombineGVCFs tool and jointly genotyped using the GenotypeGVCFs tool from GATK.

The raw SNVs and INDEls were hard-filtered using the following criteria. Filter settings for SNVs include mapping quality (< 40), variant quality score (< 30), quality by depth (< 2), strand odds ratio (> 3), Fisher strand bias (> 60), mapping quality rank-sum test (< −12.5) and read position rank-sum test (< −8). Filter settings for INDELs include quality by depth (< 2), variant quality score (< 30), Fisher strand bias (> 200) and read position rank-sum test (< −20). Very low coverage regions with a read depth < 10 and a genotype quality < 20 were set to missing on a per-sample basis. Monomorphic (no alternative alleles were called for any of the samples) and multiallelic (all sites at which only alternative alleles are called) variants, clusters of three or more SNPs called in a 10-base pair window, and variants in repetitive regions identified by TandemRepeatFinder^50^ (version 4.09.1) were removed. In addition, we removed SNVs that are within 10 bp of INDELs and allowed only one variant within a cluster of INDELs separated by 5 or fewer base pairs to pass. The remaining variants surviving the filtering process were exported in table format using the GATK VariantsToTable tool. SNVs and INDEls were annotated using the SnpEff tool^51^ (version 5.0).

### Characterization of copy number variation (CNV)

We characterized CNV for each open reading frame (ORF) across eight nuclear chromosomes. The distribution of read depth within each ORF was computed using BEDTools (version 2.31.0)^52^. This distribution was then transformed into a copy number distribution by dividing the read depth by the average read depth across all diploid chromosomes, which was then multiplied by two. Amplified or deleted regions within an ORF can be identified by analyzing the number and position of peaks within the distribution. A normal diploid ORF should display a single peak at the position of two copies, whereas amplifications or deletions result in shifts in peak positions or the emergence of extra peaks. All peaks were identified from smooth Gaussian kernels fitted to the distribution. For each peak, the percentage contribution to the overall distribution was calculated by dividing the area under the peak (boundaries determined as midpoints between this peak and nearest peaks) by the length of the ORF. Small peaks that contributed less than 20% were deemed data noise and filtered out. The remaining peak contributions were then renormalized.

Each CNV was characterized by two variables: a categorical one indicating the type of CNV (normal, full or partial amplification, and full or partial deletion), and a continuous one quantifying the copy number of the amplified or deleted regions. When only one peak remains, the CNV type was categorized as “normal” if the peak centers at two copies, “full amplification” if it centers at a copy number greater than two, or “full deletion” if it centers at a copy number less than two. In cases where two peaks remain and one of them centers at two copies, the CNV type was categorized as “partial amplification” if the other peak centers at a copy number greater than two, or “partial deletion” if it centers at a copy number less than two. If more than two peaks remain and one of them centers at two copies, the amplified or deleted region was determined by the peak with the largest percent contribution among all peaks whose copy number is not two. If two or more peaks remain but none of them center at two copies, both variables were determined based on the location of the largest peak. In the extreme scenario where all peak contributions were below 20% before filtering, these unfiltered small peaks were grouped. The quantitative copy number of this grouped peak was calculated as a weighted average of the copy numbers of these unfiltered peaks.

### Identification of *C. parapsilosis* isolates with aneuploidy

Aneuploidy was identified through the presence of multiple peaks in the distribution of average ORF (open reading frame) coverage across the entire genome. Among the 219 isolates, 15 exhibited two peaks and none exhibited more than two. In isolates with two peaks, the center of the major peak corresponded to the average read depth mapped to normal diploid chromosomes, while the minor peak represented chromosome(s) with potential aneuploidy. The copy number of chromosomes with potential aneuploidy was calculated as the ratio of the minor peak’s read depth to the major peak’s, which was then multiplied by two. We identified 12 isolates with aneuploidy that met the criteria of the copy number ≥ 2.5 or ≤ 1.5.

### Identification of inverted repeats

An inverted repeat consists of two arms of similar DNA sequences with one single stranded sequence followed downstream by its reverse complement. The inverted repeats in the *C. parapsilosis* reference genome CDC317 were identified using Inverted Repeats Finder (version 3.08). The input parameters include Match = 2, Mismatch = 3, Delta = 5, PM = 80, PI = 10, Minscore = 20, MaxLength = 100000, MaxLoop = 1000000.

### Feature encoding and merging

SNVs, INDELs, and CNV types are categorical variables and were encoded by one-hot encoding. Specifically, each SNV or INDEL was encoded by two dummy variables: a binary variable indicating whether this variant is a heterozygous allele and a binary variable indicating whether this variant is a homozygous alternative allele. Each CNV type variable was encoded by four dummy variables, each of which indicates whether the type is “full amplification”, “partial amplification”, “full deletion”, or “partial deletion”. No transformation was needed for the quantitative CNV variable.

The total number of variables, including both dummy variables created for SNVs, INDELs, CNV types and unaltered quantitative CNV variables, is 59,881. These variables were termed “raw features”. Additionally, we merged “raw features” that shared identical values across the 219 isolates. These variants that perfectly correlate with each other across multiple genomic loci suggest linkage equilibrium. A total of 6,806 merged features were identified.

### Phylogenetic tree construction

The filtered VCF file containing genomic variants served as input for the RRHS tool^53^ (version, 1.0.0.2). This tool generated haploid FASTA sequences of equal length for each isolate (parameter -n 1 generates one sequence per isolate). Subsequently, the phylogenetic tree was constructed from these sequences using IQ-TREE 2 (version 2.0.7) ^54^. IQ-TREE 2 was executed with the following parameters: -T AUTO (automatically selected threads), -bb 1000 (ultrafast bootstrap approximation ^55^ with 1000 replicates), -m MFP+ASC (use ModelFinder to determine the best fit substitution model with ascertainment bias correction), and -alrt 1000 (1000 bootstrap replicates for SH-like approximate likelihood ration test).

### Logistic regression

We used the “Logit()” function implemented in the Python statsmodels package (version 0.14.0) to perform univariate logistic regression, i.e., the association with heteroresistance was assessed for each feature individually. Raw P values were adjusted through the Bonferroni correction. A significance threshold of 0.05 was applied to the adjusted P values.

### LASSO and phylogenetic LASSO

The conventional LASSO (Least Absolute Shrinkage and Selection Operator) method determines the coefficient vector β that associates genomic features (matrix *X*) with heteroresistance label (vector *Y*) by minimizing ||*Y* − *X*β||^2^/2N + α||β||_1_, where α is a constant that multiplies the regularization term, and || · ||_1_and || · ||_2_are L1 and L2 norm respectively. To run LASSO analysis, we called the function “LassoCV” implemented in the Python’s scikit-learn library (version 1.2.2) with CV = 5 (5-fold cross validation) and n_alphas = 1,000 (number of α along the regularization path).

The phylogenetic LASSO model is an extension of conventional LASSO by controlling for variance explained by phylogeny. The technical details of this approach have been described in the Supplementary Information of our previous study for bacterial genomics^56^. Briefly, the matrix *X* (i.e., genomic features) and vector *Y* (i.e., heteroresistance label) need to be transformed before applying conventional LASSO: *X*_new_ → (T^′^T)^−1^T^′^*X*, *Y*_new_ → (T^′^T)^−1^T^′^*Y*, where matrix T is obtained from the phylogenetic variance-covariance matrix G by Cholesky decomposition such as TT^′^ = G. The phylogenetic variance-covariance matrix was computed using the “vcv.phylo” function of the ape library^57^ (version 5.7.1).

### Machine-learning-based heteroresistance prediction

All machine learning models were developed using data from a training set and subsequently assessed against a test set. As shown above, patient may harbor multiple identical fecal and blood isolates due to repeated sampling and gut-to-blood translocation. If these identical isolates were distributed into both the training and test sets, model performance may be inflated as a result of data leakage. To avoid this potential risk, we systematically excluded duplicate isolates sharing identical genomic sequences (see the section “Average nucleotide identity (ANI) and identity between isolates” for the definition of identical isolates) within each patient. This exclusion process was performed for isolates whose patient information is available.

The Random Forest (RF) and Extreme Gradient Boosting (XGBoost) models were built using a fixed number of 1,000 trees. Other hyperparameters were tuned to maximize the F1 score through grid search in a 5-fold cross-validation framework. The Decision Tree (DT) and RF models shared three parameters, including the number of features chosen by an upstream feature selection algorithm (n_features = 2, 4, 6, 8, 10), the number of features considered when determining the optimal split (max_features = square root (n_features) and n_features), and the maximum depth of each tree (max_depth = 2, 4, 6). The RF models had an extra tunable parameter—the maximum percentage of samples each tree was trained on (max_samples = 0.5, 0.75, 1.0). In the XGBoost models, two unique parameters in addition to n_features and max_depth were optimized: the percentage of features used for building each tree (colsample_bytree = 0.5, 0.75, 1.0) and the learning rate controlling the step size at which model weights were updated (learning_rate = 0.03, 0.1, 0.3).

The implementation of logistic regression and LASSO in Python has been described above. In the case of logistic regression-based feature selection, features were ranked based on their P values (from smallest to largest) and the top “n_features” were selected. When LASSO was employed for feature selection, non-zero features were ranked based on their absolute values (from largest to smallest). In situations where the coefficients of all features are zero, we randomly selected “n_features” features from the entire set of features. When the number of non-zero features was fewer than “n_features”, all non-zero features were selected. The ENNS algorithm has been implemented previously ^41^, see code at https://github.com/KaixuYang/ENNS.

The DT and RF models were constructed using the DecisionTreeClassifier and RandomForestClassifier functions from Python’s scikit-learn package (version 1.2.2), respectively. The XGBoost models were developed using the XGBClassifier function from the Python’s XGBoost package (version 1.7.6). We used four metrics, including precision, recall, F1 score (= 2*precision*recall / (precision + recall)), and accuracy, to evaluate the performance of machine learning models.

### Phylogeny-based heteroresistance prediction

We used the PICRUSt2^58^ pipeline (version 2.5.2) to predict heteroresistance phenotype. For each train-test split, we began by performing sequence placement using the “place_seqs.py” script. This step involved placing haploid sequences from test isolates into a phylogenetic tree created from training isolates. The custom input files provided to the script “place_seqs.py” included aligned haploid sequences of training isolates (generated by RRHS^53^, version 1.0.0.2), a phylogenetic tree of the training isolates and a model file specifying optimal tree parameters (generated by RAxML-NG^59^, version 1.2.0), as well as a hidden Markov model of aligned training sequences (generated using hmmbuild from hmmer.org, version 3.1b2). The output of the “place_seqs.py” script is a tree with placed test isolates. With this tree, we next ran hidden-state prediction using the “hsp.py” script to infer their heteroresistance phenotype.

### Data visualization

The data obtained from population analysis profile (PAP), time-kill, and antifungal susceptibility assays were graphed by GraphPad prism (San Diego, version 9). All other data were generated and visualized using Python’s matplotlib package (version 3.7.1). All figures underwent further editing in Adobe Illustrator (version 2021). Box plots, swarm plots, and heatmaps were generated using the corresponding functions within Python’s seaborn package (version 0.12.2). In all boxplots, the central line represents the median, the box limits denote the first and third quartiles respectively, and the whiskers extend to the smallest and largest values or at most to 1.5× the interquartile range, whichever is smaller.

### Other computational tools

In general, Python version 3.8.5 and R version 4.3.1 were used in this study for computational analyses. Moran’s I test for phylogenetic correlation was performed using “Moran.I” function from the “ape” package (version 5.7.1). Phylogenetic trees were plotted and annotated using the package “ggtree” (version 3.8.0). Fisher’s exact test and paired-sample *t*-test were performed using “fisher_exact” and “ttest_rel” from the SciPy (version 1.10.1) stats module respectively.

## Data Availability Statement

The data that support the findings of this study are available from the corresponding authors upon request. All sequencing data generated in this study will be deposited in the Sequence Read Archive upon acceptance.

## Code Availability Statement

Codes of this study are available at: https://github.com/liaochen1988/EchinocandinHeteroresistance.git

## Acknowledgements

We thank E. Pamer and K. Wolfe for discussions, and E. Zuniga for sequencing assistance. We thank D. Andes for providing clinical *C. parapsilosis* isolates. This work was supported by the National Key Research and Development Program of China 2021YFA0911300 (B.Z.), National Institutes of Health (NIH) grant nos. R01 AI093808 (T.M.H.), R21 AI105617 (T.M.H.), R21 AI156157 (T.M.H.), U19 AI158080 (D.S.W.), R01s AI141883 and AI148661 (D.S.W), U01 AI124275 (J.B.X.), R01 AI137269 (J.B.X. and Y.T.), an Investigator in the Pathogenesis of Infectious Diseases Award from the Burroughs Wellcome Fund (T.M.H. and D.S.W.), the Ludwig Center for Cancer Immunotherapy (T.M.H.), the Susan and Peter Solomon Divisional Genomics Program (T.M.H.), Deutsche Forschungsgemeinschaft (DFG, German Research Foundation) grant no. RO-5328/1-2 (T.R.), Science Foundation Ireland grant nos. 19/FFP/6668 (G.B.), 18/CRT/6214 (G.B.), and all authors from Memorial Sloan Kettering Cancer Center were supported by NIH P30 CA008748.

## Author contributions

The study was conceived by B.Z., C.L., S.J., D.S.W. and T.M.H.. MSKCC Clinical data were collected and analyzed by Y.T.(MSKCC) and B.Z.. *C. parapsilosis* isolates from MSKCC were collected and processed by B.Z., T.R., M.G., E.M., and N.E.B.. Isolates from China were maintained and processed by Y.T., T.S., and L. Z.. Isolates from Europe were collected and processed by S.B., G.B., and O. B.. Antifungal phenotypic assays were performed by S.J. and Y.N.. Whole genome sequencing analyses were done by C.L. and Y.T.(SIAT). Association analyses and machine learning modeling were done by C.L., Y.T.(SIAT), and J.B.X.. The manuscript was written by B.Z., C.L., S.J., D.S.W. and T.M.H.. All co-authors reviewed and edited the manuscript.

## Extended Data Figures

**Extended Data Fig. 1.**
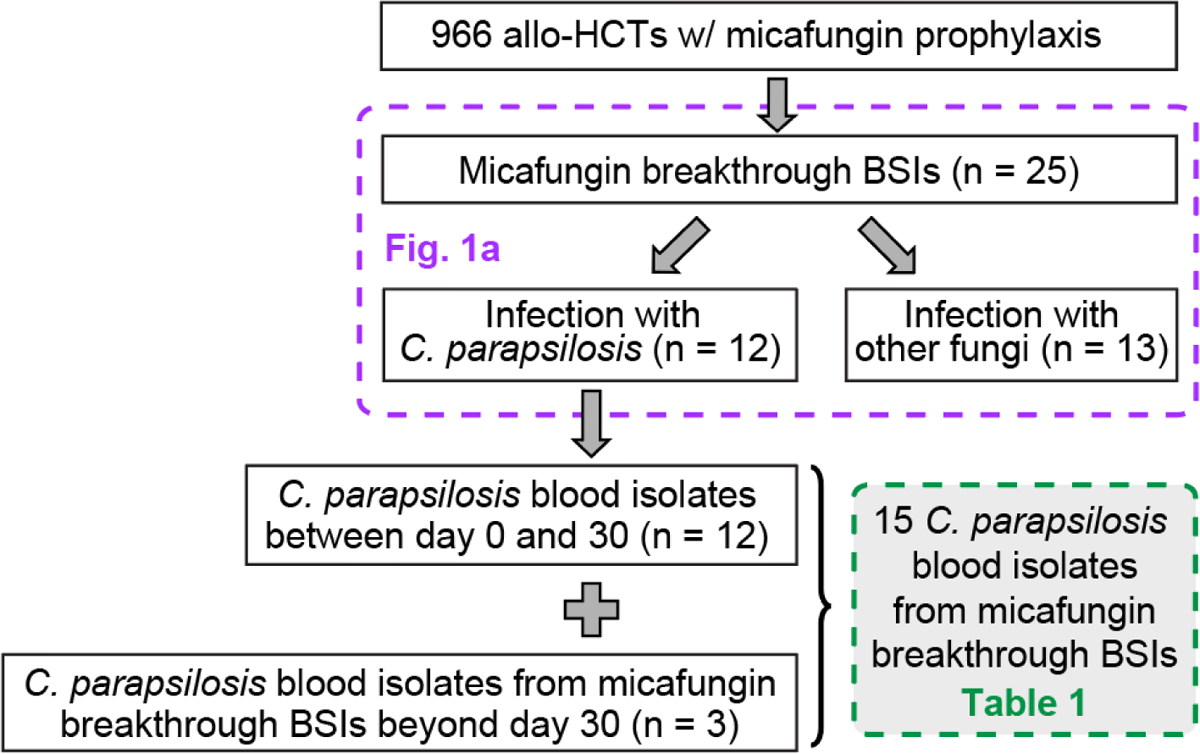
Micafungin breakthrough infections in allo-HCT recipients in MSKCC from Jan. 2016 to Dec. 2020. The dashed-line boxes indicate the patients or fungal isolates analyzed in respective figure panels or tables.

**Extended Data Fig. 2.**
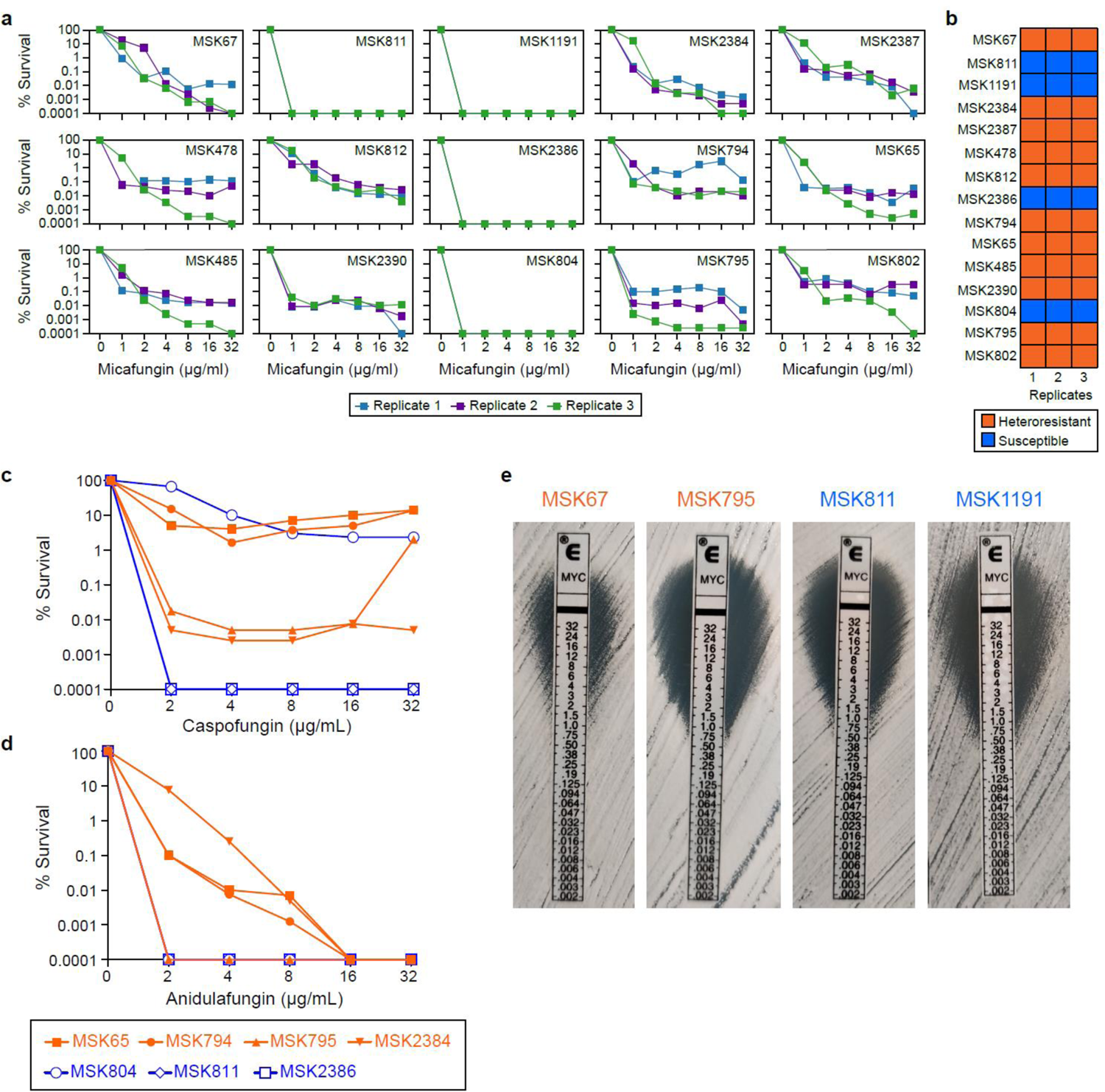
Antifungal susceptibility of *C. parapsilosis* blood isolates to echinocandin drugs. (a, b) Reproducibility of the PAP method in determining micafungin heteroresistance: a, three repeats of PAP assays of the 15 *C*. *parapsilosis* blood isolates in Table 1. b, micafungin susceptibility phenotypes (susceptible or heteroresistant) classified by the PAP curves in panel a. (c, d) Antifungal susceptibility of *C. parapsilosis* isolates to (c) caspofungin and (d) anidulafungin. Isolates MSK65, MSK794, MSK795, and MSK2384 are heteroresistant to micafungin (colored in orange), while the isolates MSK804, MSK811, and MSK2386 are susceptible to micafungin (colored in blue). Among these isolates, the phenotypes of caspofungin and anidulafungin heteroresistance were generally consistent with their micafungin heteroresistance phenotype, with exceptions of MSK795 (heteroresistant to micafungin and caspofungin, susceptible to anidulafungin) and MSK804 (susceptible to micafungin and anidulafungin, heteroresistant to caspofungin). (e) Etest results of four additional *C. parapsilosis* blood isolates. MSK67 and MSK795 are micafungin-heteroresistant (orange font), MSK811 and MSK1191 are micafungin susceptible (blue font). Notably, both heteroresistant isolate (MSK795) and susceptible isolate (MSK811) could have clean inhibition zone around the Etest strip. Again, these observations showed the challenge of using Etest to differentiate micafungin-heteroresistant and micafungin-susceptible isolates.

**Extended Data Fig. 3.**
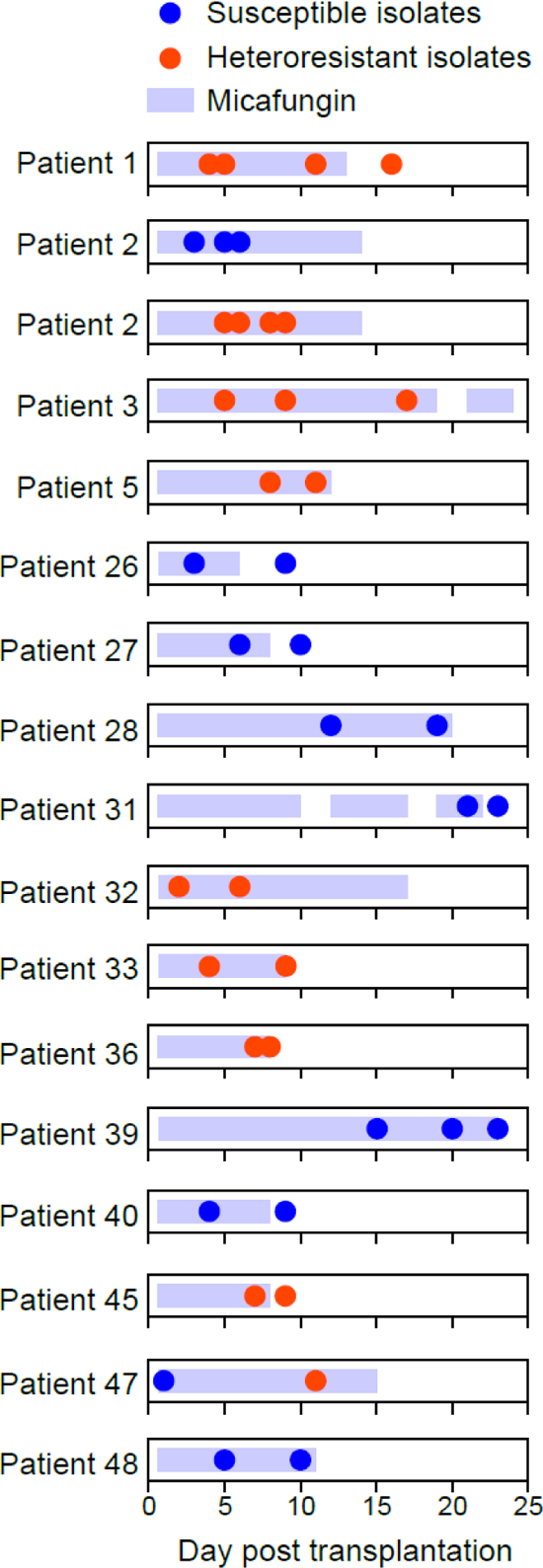
Tracking the stability of micafungin susceptibility phenotypes of identical *C. parapsilosis* fecal isolates within patients. Each filled circle represents a fecal isolate and all isolates shown in each row are identical. To simplify presentation, identical isolates collected on the same day have been consolidated into a single, filled circle. The susceptible and heteroresistant isolates obtained from Patient 2 were displayed at two separate rows.

**Extended Data Fig. 4.**
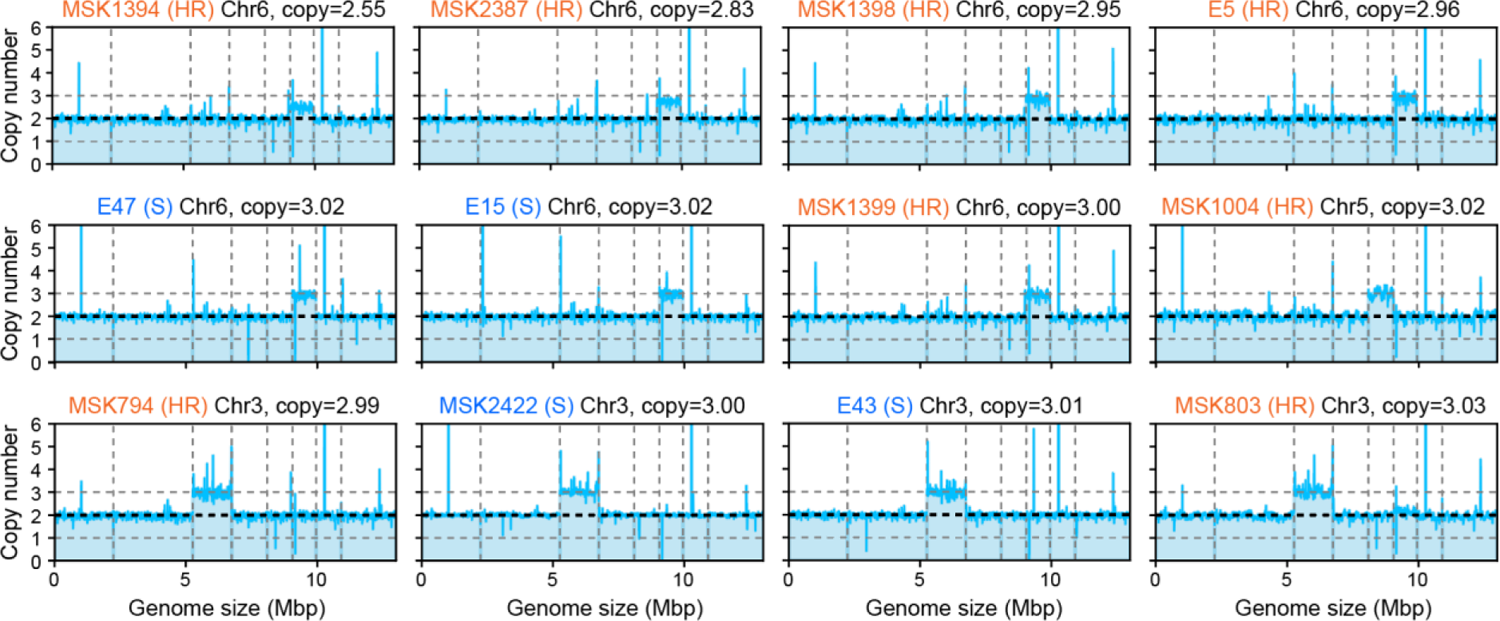
Twelve *C. parapsilosis* isolates with aneuploidy. The mean copy number of consecutive fixed-size 5kb windows over the entire genome were plotted for the isolates with aneuploidy. The mean copy number was computed as the mean read depth across each 5kb window divided by that averaged over normal diploid chromosomes. Chr: chromosome, S: susceptible, HR: heteroresistant.

**Extended Data Fig. 5.**
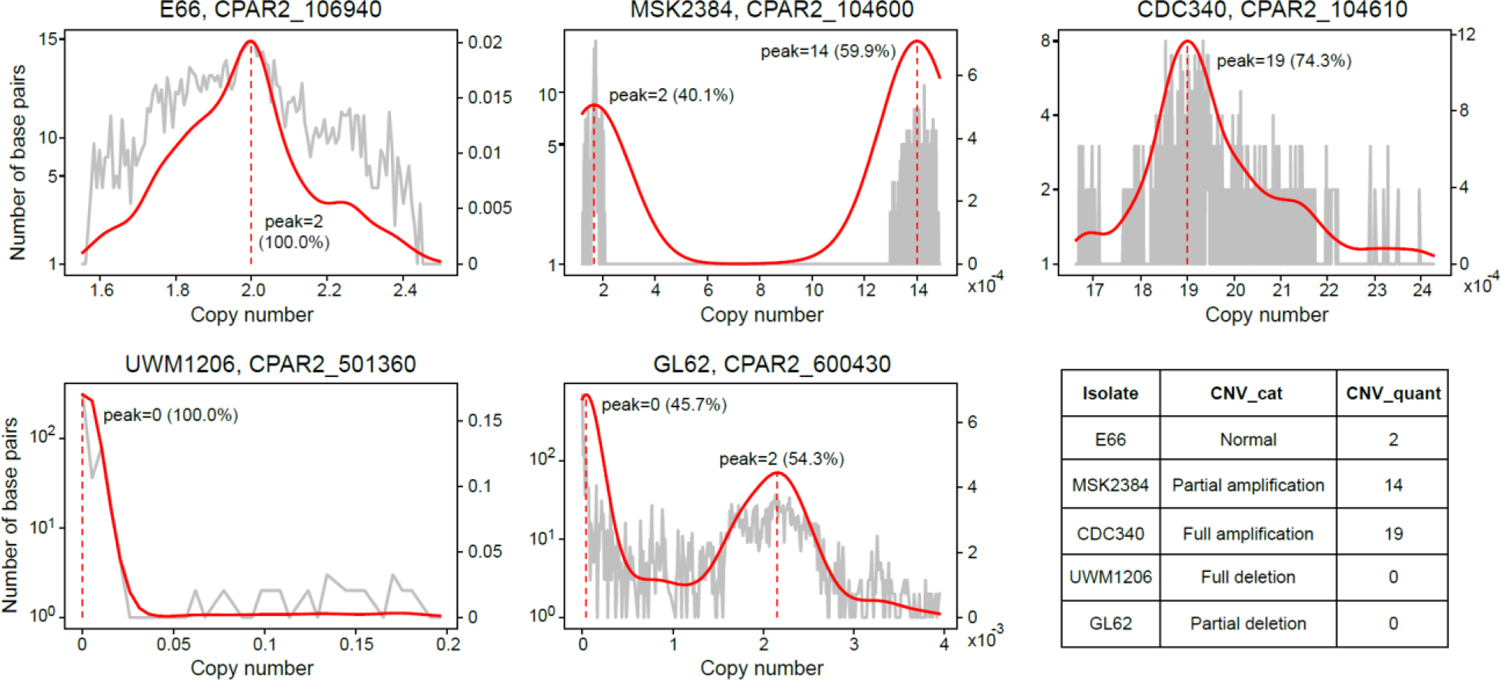
Different types of copy number variations (CNVs). To illustrate our approach for characterizing CNVs, we have provided five examples of open reading frames (ORFs) with distinct CNV types. In each panel, the light gray curve represents distribution of read depth of each position within an ORF. The solid red line fits a Gaussian kernel to this distribution. The vertical dashed red line indicates the peak center of the fitted distribution. The percentage contribution of each peak to the entire distribution is provided in the panel. The feature values of these ORFs used in following analyses (e.g., statistical association, machine learning) are shown in the table at the bottom right. Each ORF is described by a categorical feature (CNV_cat) and a quantitative feature (CNV_quant). CNV_cat specifies the CNV types (normal diploid ORF, partial amplification, full amplification, full deletion, partial deletion), while CNV_quant specifies the copy number of the amplified or deleted regions (the value of CNV_quant is 2 in the absence of amplification and deletion).

**Extended Data Fig. 6.**
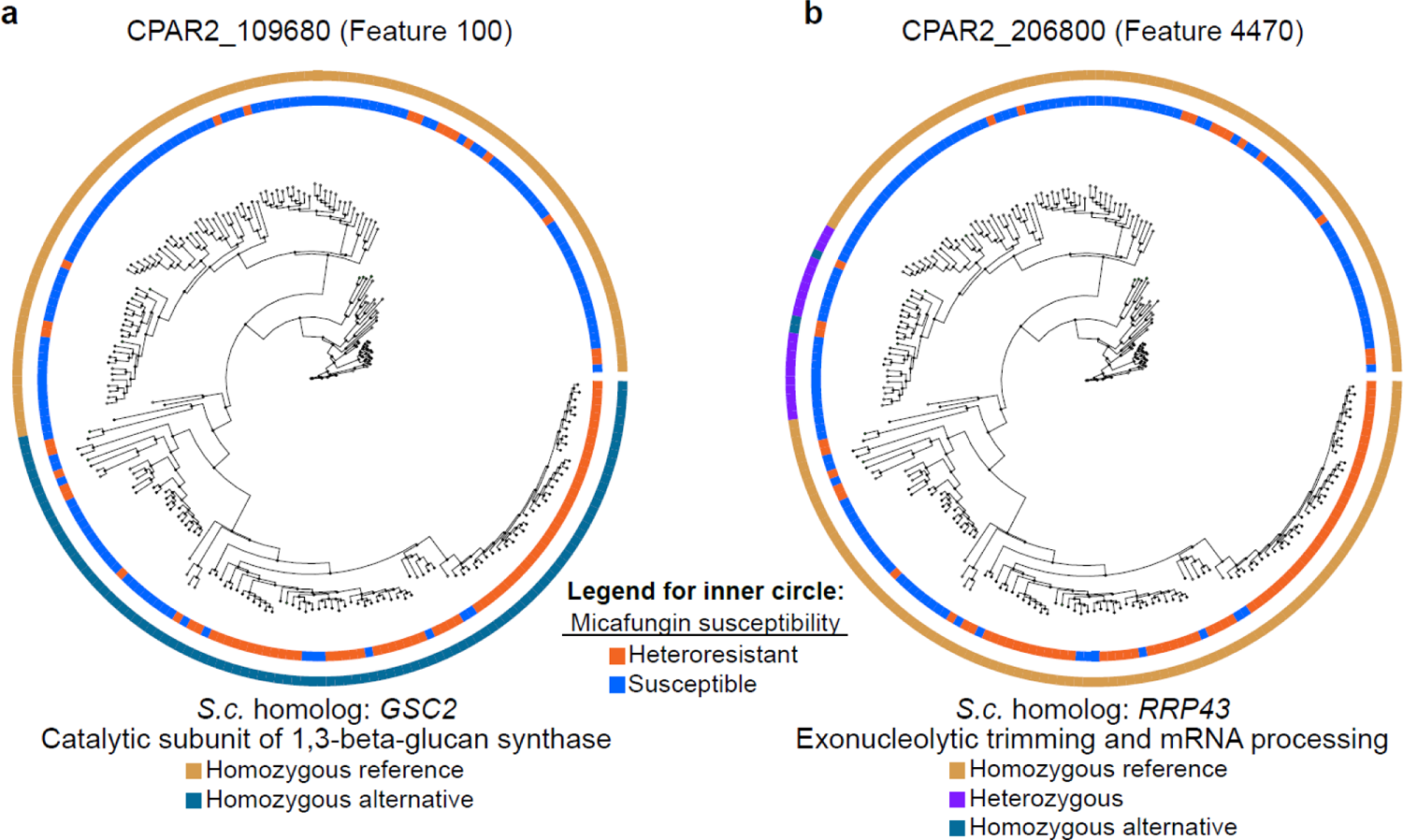
Distributions of Feature 100 (a) and 4470 (b) across the phylogenetic tree. Both features are single nucleotide variants. The biological functions of the open reading frames to which they belong are shown. *Saccharomyces cerevisiae*, *S.c*.

**Extended Data Fig. 7.**
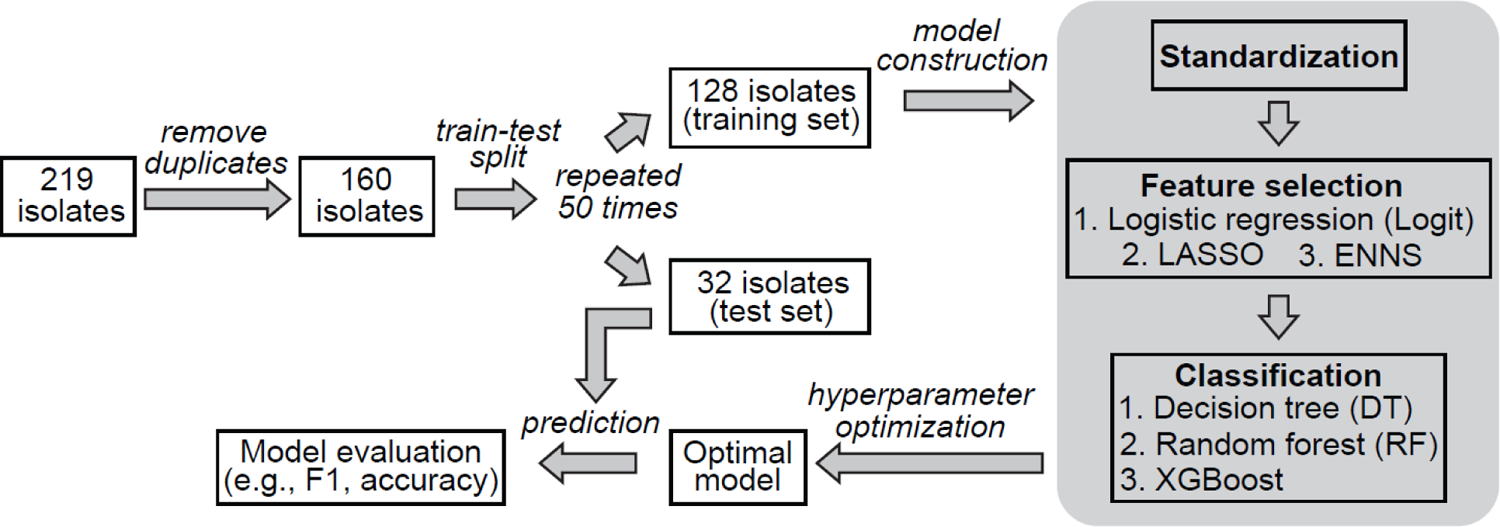
Schematic diagram of machine learning framework in predicting micafungin susceptibility phenotype. LASSO: Least Absolute Shrinkage and Selection Operator; ENNS: Ensemble Neural Network Selection; XGBoost: Extreme Gradient Boosting.

**Extended Data Fig. 8.**
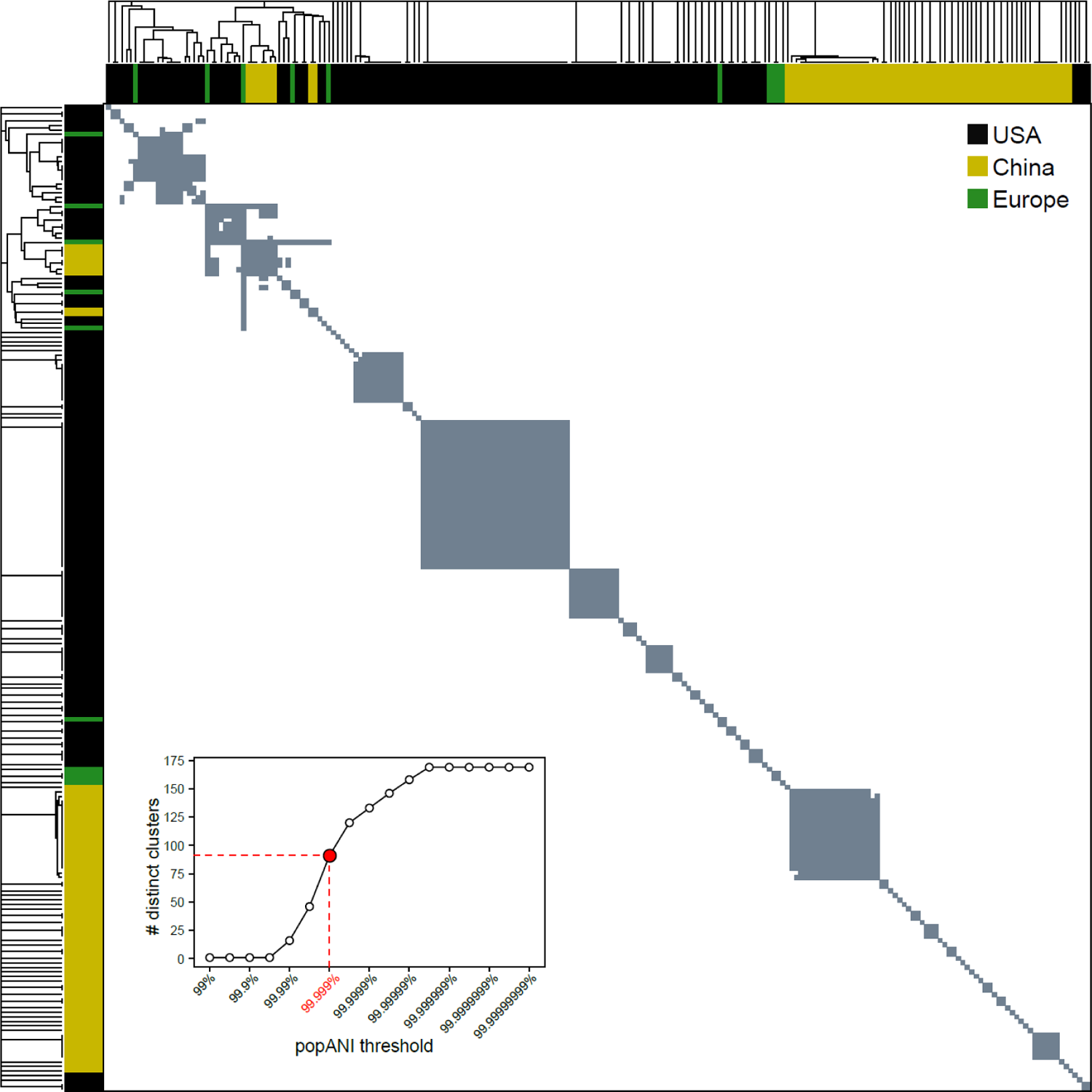
Hierarchical clustering of pairwise popANI (population average nucleotide identity) values across all 219 *C. parapsilosis* isolates. Each row or column represents an individual isolate. A gray box indicates a popANI value of at least 99.999% between the respective row and column isolates, while a white box indicates a value below this cutoff. Using the cutoff, we identified 91 clusters of varying sizes (*inset*, red circle; see Methods). For each cluster that contains at least two isolates, any pair of isolates within this cluster has a popANI value of at least 99.999%, indicating that these isolates are all identical to each other. The *inset* curve also shows the impact of the popANI threshold on the number of identified clusters. The detailed pairwise popANI values are available in Supplementary Table 9.

**Extended Data Fig. 9.**
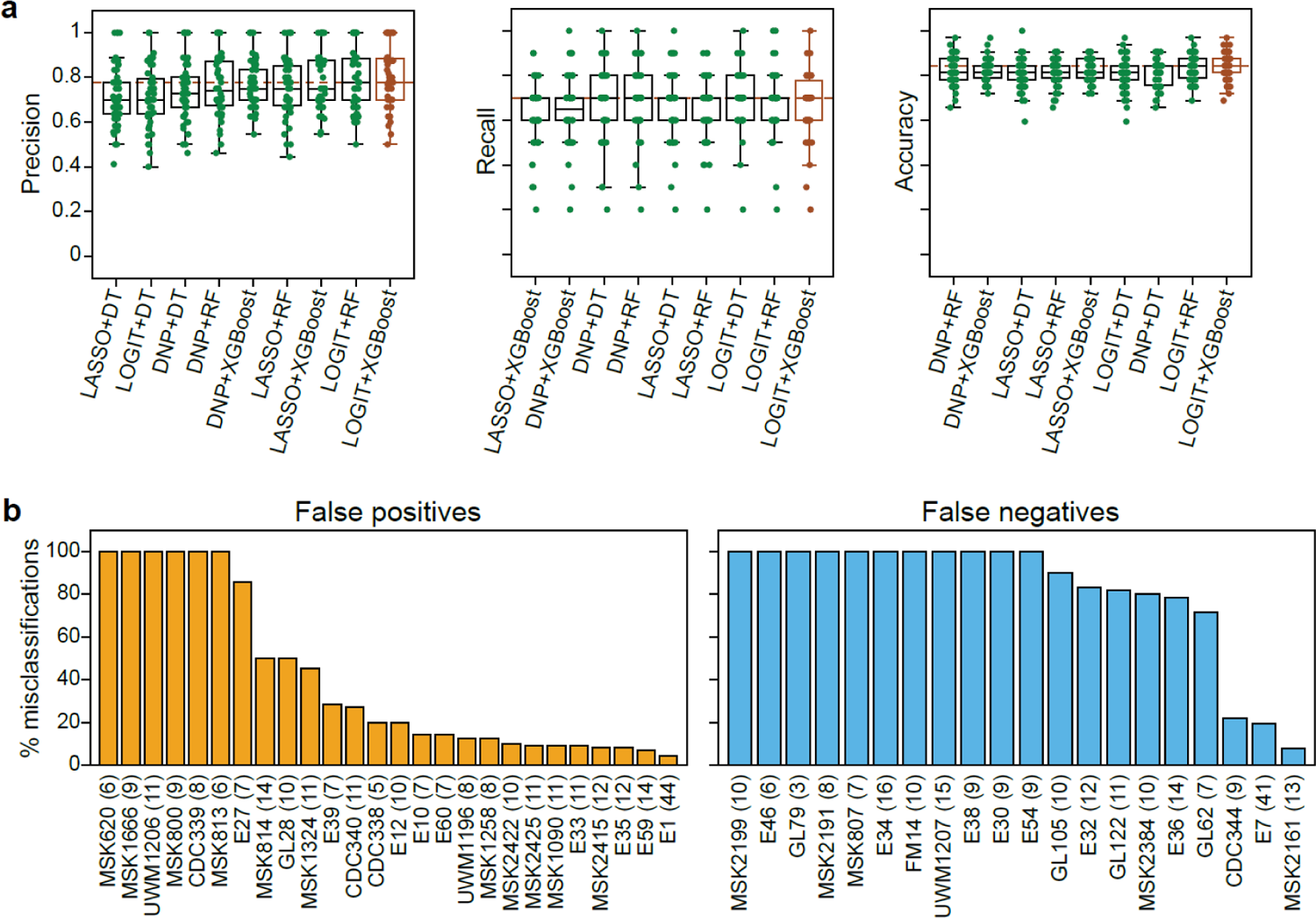
Performance of machine learning models. **(a)** Precision, recall, and accuracy scores of machine learning models constructed with different combinations of feature selection algorithms (LOGIT, LASSO, ENNS) and classifiers (DT, RF, XGBoost). In all boxplots, each dot represents a single train-test split, which was randomly repeated 50 times. The models were ranked based on their median scores in the ascending order. LOGIT+XGBoost always achieved the highest median score. LOGIT, Logistic regression; LASSO, Least Absolute Shrinkage and Selection Operator; ENNS, Ensemble Neural Network Selection; DT, Decision Tree; RF, Random Forest; XGBoost, Extreme Gradient Boosting. **(b)** Misclassification (both false-positive and negative predictions) percentage for all isolates misclassified at least once among 50 random train-test splits. The number within parenthesis following each isolate identifier indicates how many times the isolate was included in the test set.

**Extended Data Fig. 10.**
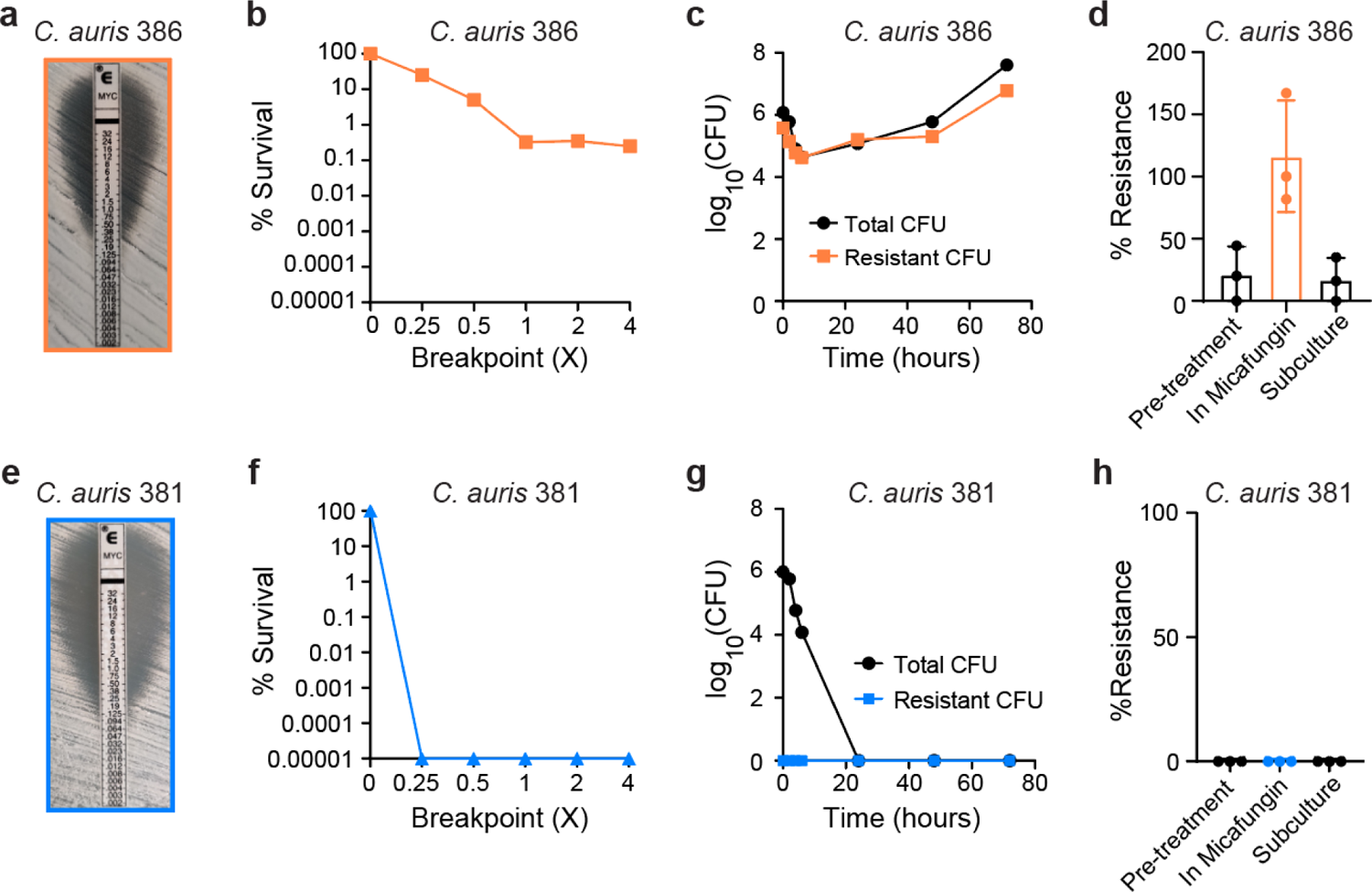
Micafungin heteroresistance in Candida auris clinical isolates. *C. auris* isolate **(a – d**) 386 and **(e – h)** 381 were obtained from CDC/FDA AR Bank. **(a**, **e)** Micafungin E-test results for *C. auris* isolates **(a)** 386 and **(e)** 381. **(b**, **f)** Micafungin population analysis profiles for *C. auris* isolates **(b)** 386 and **(f)** 381. Percent survival was calculated compared to growth on drug-free agar. **(c**, **g)** CFU dynamics of total and resistant subpopulations of *C. auris* isolates **(c)** 386 and **(g)** 381 under micafungin pressure (0.5 µg/mL). Cultures were plated at indicated time points for enumeration of total (black, circles) and resistant (colored, squares) cells. **(d, h)**, Frequency of micafungin-resistant subpopulation of *C. auris* isolates **(d)** 386 and **(h)** 381 during continuous passage. The frequency was calculated after a 24-hour growth period in YPD broth (Pre-treatment), followed by a 48-hour passage in YPD broth with 0.5 µg/mL micafungin (In micafungin), and subsequently, a passage in YPD broth without micafungin for 24 hours (Subculture). Bar height: mean; error bars: standard deviation (n=3).

## Supplementary information

**Supplementary Table 1.** Population analysis profile (PAP) data of all 219 *C. parapsilosis* isolates in this study.

**Supplementary Table 2.** Information of patients and *C. parapsilosis* isolates in this study.

**Supplementary Table 3**. Copy number variations (CNVs) of open reading frames (ORFs) identified from 219 *C. parapsilosis* isolates.

**Supplementary Table 4**. ORFs with amplification that overlap with inverted repeats of at least 100bp.

**Supplementary Table 5**. Single nucleotide variants (SNVs) and insertion/deletions (INDELs) identified from the 219 *C. parapsilosis* isolates.

**Supplementary Table 6.** Mapping between raw and merged features. Raw features of single nucleotide variants (SNVs), insertion/deletions (INDELs), and copy number variations (CNVs) were merged if they held identical values across all 219 isolates. The identifiers of raw SNV and INDEL features follow the pattern: “Chromosome ID:Position:SNP (or INDEL):Reference allele:Alternative allele=X”, where X=1, 2 denotes heterozygous and homozygous alternative alleles respectively. For categorial CNVs, the raw feature identifier follows the pattern “CNV_cat [open reading frame identifier]=X”, where X = 1, 2, 3, 4 represent full amplification, partial amplification, full deletion, and partial deletion respectively. The identifiers of raw quantitative CNV features has the format “CNV_quant [open reading frame identifier]”.

**Supplementary Table 7.** Logistic regression analysis of the correlation between genomic features and micafungin heteroresistance. The logistic regression coefficients, along with their lower and upper confidence intervals (CIs) and adjusted P values (P_adj._), are provided. Here, the genomic features refer to the merged features in Supplementary Table 5. Only features with adjusted P values < 0.05 are included in the table.

**Supplementary Table 8**. LASSO (Least Absolute Shrinkage and Selection Operator) and phylogenetic LASSO analyses of the correlation between genomic features and micafungin heteroresistance. Features with non-zero coefficients in at least one analysis were included in the table.

**Supplementary Table 9.** Clustering of *C. parapsilosis* isolates based on popANI (population average nucleotide identity). In each cluster, all isolates are identical to each other (i.e., all pairwise popANI values ≥ 99.999%). The column “Used” indicates which isolates have been included in the development of machine learning models.

## Reference

1 Antimicrobial Resistance, C. Global burden of bacterial antimicrobial resistance in 2019: a systematic analysis. Lancet 399, 629–655, doi:10.1016/S0140-6736(21)02724-0 (2022).

2 Berman, J. & Krysan, D. J. Drug resistance and tolerance in fungi. Nat Rev Microbiol 18, 319–331, doi:10.1038/s41579-019-0322-2 (2020).

3 Andersson, D. I., Nicoloff, H. & Hjort, K. Mechanisms and clinical relevance of bacterial heteroresistance. Nat Rev Microbiol 17, 479–496, doi:10.1038/s41579-019-0218-1 (2019).

4 El-Halfawy, O. M. & Valvano, M. A. Antimicrobial heteroresistance: an emerging field in need of clarity. Clin Microbiol Rev 28, 191–207, doi:10.1128/CMR.00058-14 (2015).

5 Band, V. I. & Weiss, D. S. Heteroresistance: A cause of unexplained antibiotic treatment failure? PLoS Pathog 15, e1007726, doi:10.1371/journal.ppat.1007726 (2019).

6 Mondon, P. et al. Heteroresistance to fluconazole and voriconazole in Cryptococcus neoformans. Antimicrob Agents Chemother 43, 1856–1861, doi:10.1128/AAC.43.8.1856 (1999).

7 Sionov, E., Chang, Y. C., Garraffo, H. M. & Kwon-Chung, K. J. Heteroresistance to fluconazole in Cryptococcus neoformans is intrinsic and associated with virulence. Antimicrob Agents Chemother 53, 2804–2815, doi:10.1128/AAC.00295-09 (2009).

8 Ben-Ami, R. et al. Heteroresistance to Fluconazole Is a Continuously Distributed Phenotype among Candida glabrata Clinical Strains Associated with In Vivo Persistence. mBio 7, doi:10.1128/mBio.00655-16 (2016).

9 Band, V. I. et al. Antibiotic failure mediated by a resistant subpopulation in Enterobacter cloacae. Nat Microbiol 1, 16053, doi:10.1038/nmicrobiol.2016.53 (2016).

10 Xiong, Y. et al. Impact of Carbapenem Heteroresistance Among Multidrug-Resistant ESBL/AmpC-Producing Klebsiella pneumoniae Clinical Isolates on Antibiotic Treatment in Experimentally Infected Mice. Infect Drug Resist 14, 5639–5650, doi:10.2147/IDR.S340652 (2021).

11 Bert, F. et al. Prevalence, molecular epidemiology, and clinical significance of heterogeneous glycopeptide-intermediate Staphylococcus aureus in liver transplant recipients. J Clin Microbiol 41, 5147–5152, doi:10.1128/JCM.41.11.5147-5152.2003 (2003).

12 Sieradzki, K., Roberts, R. B., Serur, D., Hargrave, J. & Tomasz, A. Heterogeneously vancomycin-resistant Staphylococcus epidermidis strain causing recurrent peritonitis in a dialysis patient during vancomycin therapy. J Clin Microbiol 37, 39–44, doi:10.1128/JCM.37.1.39-44.1999 (1999).

13 Fisher, B. T. et al. Effect of Caspofungin vs Fluconazole Prophylaxis on Invasive Fungal Disease Among Children and Young Adults With Acute Myeloid Leukemia: A Randomized Clinical Trial. JAMA 322, 1673–1681, doi:10.1001/jama.2019.15702 (2019).

14 Walsh, T. J. et al. Caspofungin versus liposomal amphotericin B for empirical antifungal therapy in patients with persistent fever and neutropenia. N Engl J Med 351, 1391–1402, doi:10.1056/NEJMoa040446 (2004).

15 Slavin, M. A. et al. Efficacy and safety of fluconazole prophylaxis for fungal infections after marrow transplantation--a prospective, randomized, double-blind study. J Infect Dis 171, 1545–1552, doi:10.1093/infdis/171.6.1545 (1995).

16 Goodman, J. L. et al. A controlled trial of fluconazole to prevent fungal infections in patients undergoing bone marrow transplantation. N Engl J Med 326, 845–851, doi:10.1056/NEJM199203263261301 (1992).

17 Epstein, D. J., Seo, S. K., Brown, J. M. & Papanicolaou, G. A. Echinocandin prophylaxis in patients undergoing haematopoietic cell transplantation and other treatments for haematological malignancies. J Antimicrob Chemother 73, i60–i72, doi:10.1093/jac/dkx450 (2018).

18 Douglas, C. M. et al. Identification of the FKS1 gene of Candida albicans as the essential target of 1,3-beta-D-glucan synthase inhibitors. Antimicrob Agents Chemother 41, 2471–2479, doi:10.1128/AAC.41.11.2471 (1997).

19 Toth, R. et al. Candida parapsilosis: from Genes to the Bedside. Clin Microbiol Rev 32, doi:10.1128/CMR.00111-18 (2019).

20 Daneshnia, F. et al. Worldwide emergence of fluconazole-resistant Candida parapsilosis: current framework and future research roadmap. Lancet Microbe 4, e470–e480, doi:10.1016/S2666-5247(23)00067-8 (2023).

21 Trevijano-Contador, N. et al. Global Emergence of Resistance to Fluconazole and Voriconazole in Candida parapsilosis in Tertiary Hospitals in Spain During the COVID-19 Pandemic. Open Forum Infect Dis 9, ofac605, doi:10.1093/ofid/ofac605 (2022).

22 Thomaz, D. Y. et al. A Brazilian Inter-Hospital Candidemia Outbreak Caused by Fluconazole-Resistant Candida parapsilosis in the COVID-19 Era. J Fungi (Basel) 8, doi:10.3390/jof8020100 (2022).

23 Fekkar, A. et al. Hospital outbreak of fluconazole-resistant Candida parapsilosis: arguments for clonal transmission and long-term persistence. Antimicrob Agents Chemother 95, doi:10.1128/AAC.02036-20 (2023).

24 Zhai, B. et al. High-resolution mycobiota analysis reveals dynamic intestinal translocation preceding invasive candidiasis. Nat Med 26, 59–64, doi:10.1038/s41591-019-0709-7 (2020).

25 Garcia-Effron, G., Katiyar, S. K., Park, S., Edlind, T. D. & Perlin, D. S. A naturally occurring proline-to-alanine amino acid change in Fks1p in Candida parapsilosis, Candida orthopsilosis, and Candida metapsilosis accounts for reduced echinocandin susceptibility. Antimicrob Agents Chemother 52, 2305–2312, doi:10.1128/AAC.00262-08 (2008).

26 Pfaller, M. A., Moet, G. J., Messer, S. A., Jones, R. N. & Castanheira, M. Geographic variations in species distribution and echinocandin and azole antifungal resistance rates among Candida bloodstream infection isolates: report from the SENTRY Antimicrobial Surveillance Program (2008 to 2009). J Clin Microbiol 49, 396–399, doi:10.1128/JCM.01398-10 (2011).

27 Ning, Y. et al. Decreased echinocandin susceptibility in Candida parapsilosis causing candidemia and emergence of a pan-echinocandin resistant case in China. Emerg Microbes Infect 12, 2153086, doi:10.1080/22221751.2022.2153086 (2023).

28 Wang, E. et al. The ever-evolving landscape of candidaemia in patients with acute leukaemia: non-susceptibility to caspofungin and multidrug resistance are associated with increased mortality. J Antimicrob Chemother 70, 2362–2368, doi:10.1093/jac/dkv087 (2015).

29 Lortholary, O. et al. Recent exposure to caspofungin or fluconazole influences the epidemiology of candidemia: a prospective multicenter study involving 2,441 patients. Antimicrob Agents Chemother 55, 532–538, doi:10.1128/AAC.01128-10 (2011).

30 Rolling, T. et al. Haematopoietic cell transplantation outcomes are linked to intestinal mycobiota dynamics and an expansion of Candida parapsilosis complex species. Nat Microbiol 6, 1505–1515, doi:10.1038/s41564-021-00989-7 (2021).

31 Giske, C. G., Turnidge, J., Canton, R., Kahlmeter, G. & Committee, E. S. Update from the European Committee on Antimicrobial Susceptibility Testing (EUCAST). J Clin Microbiol 60, e0027621, doi:10.1128/JCM.00276-21 (2022).

32 Lewis, K. Persister cells, dormancy and infectious disease. Nature Reviews Microbiology 5, 48–56, doi:10.1038/nrmicro1557 (2007).

33 Olm, M. R. et al. inStrain profiles population microdiversity from metagenomic data and sensitively detects shared microbial strains. Nat Biotechnol 39, 727–736, doi:10.1038/s41587-020-00797-0 (2021).

34 Sionov, E., Chang, Y. C. & Kwon-Chung, K. J. Azole heteroresistance in Cryptococcus neoformans: emergence of resistant clones with chromosomal disomy in the mouse brain during fluconazole treatment. Antimicrob Agents Chemother 57, 5127–5130, doi:10.1128/AAC.00694-13 (2013).

35 Stone, N. R. et al. Dynamic ploidy changes drive fluconazole resistance in human cryptococcal meningitis. J Clin Invest 129, 999–1014, doi:10.1172/JCI124516 (2019).

36 Yang, F. et al. Aneuploidy Underlies Tolerance and Cross-Tolerance to Drugs in Candida parapsilosis. Microbiol Spectr 9, e0050821, doi:10.1128/Spectrum.00508-21 (2021).

37 Todd, R. T. & Selmecki, A. Expandable and reversible copy number amplification drives rapid adaptation to antifungal drugs. Elife 9, e58349 (2020).

38 Zhai, B. et al. High-resolution mycobiota analysis reveals dynamic intestinal translocation preceding invasive candidiasis. Nature medicine 26, 59–64 (2020).

39 Lai, M. H., Silverman, S. J., Gaughran, J. P. & Kirsch, D. R. Multiple copies of PBS2, MHP1 or LRE1 produce glucanase resistance and other cell wall effects in Saccharomyces cerevisiae. Yeast 13, 199–213, doi:10.1002/(SICI)1097-0061(19970315)13:3<199::AID-YEA76>3.0.CO;2-Z (1997).

40 Oh, S.-H. et al. Agglutinin-like sequence (ALS) genes in the Candida parapsilosis species complex: blurring the boundaries between gene families that encode cell-wall proteins. Frontiers in microbiology 10, 781 (2019).

41 Yang, K. & Maiti, T. ENNS: Variable Selection, Regression, Classification and Deep Neural Network for High-Dimensional Data. arXiv preprint arXiv:2107.03430 (2021).

42 Healey, K. R. et al. Prevalent mutator genotype identified in fungal pathogen Candida glabrata promotes multi-drug resistance. Nat Commun 7, 11128, doi:10.1038/ncomms11128 (2016).

43 Ostrowsky, B. et al. Candida auris Isolates Resistant to Three Classes of Antifungal Medications - New York, 2019. MMWR Morb Mortal Wkly Rep 69, 6–9, doi:10.15585/mmwr.mm6901a2 (2020).

44 Park, J. Y., Bradley, N., Brooks, S., Burney, S. & Wassner, C. Management of Patients with Candida auris Fungemia at Community Hospital, Brooklyn, New York, USA, 2016-2018(1). Emerg Infect Dis 25, 601–602, doi:10.3201/eid2503.180927 (2019).

45 Kilburn, S. et al. Antifungal Resistance Trends of Candida auris Clinical Isolates, New York-New Jersey, 2016-2020. Antimicrob Agents Chemother, aac0224221, doi:10.1128/aac.02242-21 (2022).

46 CDC. Recommendations for treatment of Candida auris infections, <https://www.cdc.gov/fungal/candida-auris/c-auris-treatment.html> (2021).

47 Taur, Y. et al. Reconstitution of the gut microbiota of antibiotic-treated patients by autologous fecal microbiota transplant. Sci Transl Med 10, doi:10.1126/scitranslmed.aap9489 (2018).

48 Kim, S. G. et al. Microbiota-derived lantibiotic restores resistance against vancomycin-resistant Enterococcus. Nature 572, 665–669, doi:10.1038/s41586-019-1501-z (2019).

49 Butler, G. et al. Evolution of pathogenicity and sexual reproduction in eight Candida genomes. Nature 459, 657–662, doi:10.1038/nature08064 (2009).

50 Benson, G. Tandem repeats finder: a program to analyze DNA sequences. Nucleic Acids Res 27, 573–580, doi:10.1093/nar/27.2.573 (1999).

51 Cingolani, P. et al. A program for annotating and predicting the effects of single nucleotide polymorphisms, SnpEff: SNPs in the genome of Drosophila melanogaster strain w1118; iso-2; iso-3. Fly (Austin) 6, 80–92, doi:10.4161/fly.19695 (2012).

52 Quinlan, A. R. & Hall, I. M. BEDTools: a flexible suite of utilities for comparing genomic features. Bioinformatics 26, 841–842, doi:10.1093/bioinformatics/btq033 (2010).

53 Lischer, H. E., Excoffier, L. & Heckel, G. Ignoring heterozygous sites biases phylogenomic estimates of divergence times: implications for the evolutionary history of microtus voles. Mol Biol Evol 31, 817–831, doi:10.1093/molbev/mst271 (2014).

54 Minh, B. Q. et al. IQ-TREE 2: New Models and Efficient Methods for Phylogenetic Inference in the Genomic Era. Mol Biol Evol 37, 1530–1534, doi:10.1093/molbev/msaa015 (2020).

55 Hoang, D. T., Chernomor, O., von Haeseler, A., Minh, B. Q. & Vinh, L. S. UFBoot2: Improving the Ultrafast Bootstrap Approximation. Mol Biol Evol 35, 518–522, doi:10.1093/molbev/msx281 (2018).

56 Yan, J. et al. Bow-tie signaling in c-di-GMP: Machine learning in a simple biochemical network. PLoS Comput Biol 13, e1005677, doi:10.1371/journal.pcbi.1005677 (2017).

57 Paradis, E., Claude, J. & Strimmer, K. APE: Analyses of Phylogenetics and Evolution in R language. Bioinformatics 20, 289–290, doi:10.1093/bioinformatics/btg412 (2004).

58 Douglas, G. M. et al. PICRUSt2 for prediction of metagenome functions. Nature biotechnology 38, 685–688 (2020).

59 Kozlov, A. M., Darriba, D., Flouri, T., Morel, B. & Stamatakis, A. RAxML-NG: a fast, scalable and user-friendly tool for maximum likelihood phylogenetic inference. Bioinformatics 35, 4453–4455 (2019).

